# Mental health symptom changes by sex or gender during COVID-19 compared to pre-pandemic: a systematic review and meta-analysis update

**DOI:** 10.64898/2026.01.22.26344371

**Authors:** Gabriel Chun Yin Sung, Yin Wu, Suiqiong Fan, Tiffany Dal Santo, Nadia P. González-Domínguez, Ying Sun, Letong Li, Kexin Li, Xiaowen Jiang, Amina Tasleem, Yutong Wang, Jill T. Boruff, Palak Desai, Brienne Tougas, Monica D’Onofrio, Ankur Krishnan, Claire Adams, Chen He, Richard S. Henry, Afra Alkan, Danielle B. Rice, Sarah Markham, Marleine Azar, Elsa-Lynn Nassar, Sophie Hu, Mara Cañedo-Ayala, Dipika Neupane, Andrea Benedetti, Brett D Thombs

## Abstract

**KEY POINTS:** *Question:* Did changes in mental health symptoms from pre-COVID-19 to during the pandemic differ by sex or gender?

*Findings:* In this systematic review and meta-analysis study of findings from 27 unique cohorts, changes from before to during the COVID-19 pandemic in mental health symptoms, including general mental health, anxiety, depression, and stress, were not statistically significantly different by sex or gender.

*Meaning:* There were not likely substantial sex and gender differences in mental health symptom changes during the COVID-19 pandemic.

**Importance:** Concerns remain about whether COVID-19 affected mental health differently across sex or gender groups. We previously reported that changes in general mental health and anxiety symptoms, but not depression or stress, worsened more for females or women than males or men, but that was based on 12 studies published up to August 2021.

**Objective:** To investigate the sex or gender differences in mental health symptoms before and during the COVID-19 pandemic with updated evidence.

**Data Sources:** MEDLINE, PsycINFO, CINAHL, EMBASE, Web of Science, China National Knowledge Infrastructure, Wanfang, medRxiv, Open Science Framework Preprints searched to August 31, 2023.

**Study Selection:** Eligible studies included change data for general mental health, anxiety symptoms, depression symptoms, or stress from pre-to during COVID-19 by sex or gender. Two independent reviewers reviewed citations for eligibility.

**Data Extraction and Synthesis:** Standardized mean differences (SMDs) were extracted for changes of continuous outcomes and proportions for dichotomous outcomes. Two independent reviewers completed data extraction and risk of bias assessment with an adapted Joanna Briggs Institute Checklist for Prevalence Studies. Data were pooled by random-effects models.

**Main Outcomes and Measures:** Difference in change of SMDs and proportions between sex or gender groups pre-COVID-19 to COVID-19.

**Results:** We included data from 27 unique cohorts (N = 102 to 18,127). Change differences by sex or gender were minimal and not statistically significant: general mental health (SMD change_women-men_ = 0.01, 95% confidence interval [CI]:-0.07 to 0.10; proportion change_women-men_ = −0.03, 95% CI: −0.08 to 0.03), anxiety (SMD change_women-men_ = 0.09, 95% CI −0.04 to 0.22; proportion change_women-men_ = −0.05, 95% CI: −0.20 to 0.11), depression (SMD change_women-men_ = 0.10, 95% CI: −0.00 to 0.20; proportion change_women-men_ = −0.13, 95% CI: −0.81 to 0.55), and stress (SMD change_women-men_ = −0.08, 95% CI −0.16 to 0.01, proportion change_women-men_ = 0.04, 95% CI −0.10 to 0.17). No studies reported eligible mental health outcomes for gender minorities.

**Conclusion and Relevance:** We found no significant sex or gender differences in mental health changes. Future research should report outcomes for gender minority groups, even if small numbers, to support evidence synthesis.

**Registration:** PROSPERO (CRD42020179703)

## INTRODUCTION

There are concerns that COVID-19 may have affected mental health among vulnerable groups, including women, disproportionately.^1–3^ Although the COVID-19 public health emergency has ended,^4^ understanding mental health impacts remains important to prepare for future public health emergencies.^5^

The most comprehensive systematic review of studies that compared pre-COVID-19 mental health symptoms and symptoms during the pandemic found that aggregate changes across groups analyzed were small (standardized mean difference [SMDs] changes pre-COVID-19 to during COVID-19 = 0.05 to 0.22); women and females were the only group with statistically significant declines across all 3 mental health categories evaluated, including general mental health (6 studies, 10,329 participants, SMD = 0.22, 95% confidence interval [CI] 0.08 to 0.35), anxiety symptoms (5 studies, 3,500 participants, SMD = 0.20, 95% CI 0.12 to 0.29), and depression symptoms (7 studies, 3,851 participants, SMD = 0.22, 95% CI 0.05 to 0.40).^1^

A sub-study of the main systematic review evaluated studies with data to directly compare mental health changes among women versus men and found significantly greater worsening for women in general mental health (3 cohorts, 15,692 participants, SMD_women-men_ = 0.15, 95% CI 0.12 to 0.18) and anxiety symptoms (4 cohorts, 4,344 participants, SMD_women-men_ = 0.15, 95% CI 0.07 to 0.22) but not depression symptoms (4 cohorts, 4,475 participants, SMD_women-men_ = 0.12, 95% CI −0.09 to 0.33) or stress (4 cohorts, 1,533 participants, SMD_women-men_ = −0.10, 95% CI −0.21 to 0.01). That analysis, however, included only evidence from searches up to August 30, 2021 and data from 12 publications.^2^

The present study updated the previous comparison. Objectives were to (1) compare changes from pre-COVID-19 to during COVID-19 on continuous general mental health, anxiety symptoms, depression symptoms, and stress using data from studies that reported data for both women and men and (2) compare changes in the proportion of women and men above study-defined cutoffs for general mental health, anxiety symptoms, depression symptoms, and stress.

## METHODS

Our main COVID-19 mental health changes systematic review was registered in PROSPERO (CRD42020179703), and a protocol was posted to the Open Science Framework prior to initiating searches (https://osf.io/96csg/). The present study is a sub-study of our main review.^1^ Results are reported per the PRISMA statement.^6^ Our methods are adopted from the main systematic review^1^ and our previous study that compared changes by sex or gender.^2^ Thus, we followed reporting guidance from the Text Recycling Research Project.^7^

### Eligibility Criteria

In our main review, we included studies on any population that compared pre-COVID-19 mental health outcomes, with at least 80% of participants assessed from January 1, 2018 to December 31, 2019 when the World Health Organization received the first report of COVID-19 from Chinese authorities to outcomes collected January 1, 2020 or later. Studies had to report data from cohorts comprising at least 90% of the same participants between pre-COVID-19 and during-COVID-19 periods or statistically account for missing data. Studies with < 100 participants were excluded for feasibility and due to their limited relative incremental value. For our sub-study, studies had to report mental health outcomes separately by sex (assignment based on external genitalia, usually at birth; e.g., female, male, intersex) or gender (socially constructed characteristics of roles and behaviours; e.g., cisgender woman, cisgender man, trans woman, trans man, non-binary).^8^

### Search Strategy

We searched Medline (Ovid), PsycINFO (Ovid), CINAHL (EBSCO), Embase (Ovid), Web of Science Core Collection: Citation Indexes, China National Knowledge Infrastructure, Wanfang, medRxiv, and Open Science Framework Preprints with a strategy designed by an experienced health sciences librarian. The China National Knowledge Infrastructure and Wanfang databases were searched using Chinese terms based on our English-language strategy. COVID-19 terms were developed in collaboration with other librarians working on the topic and updated as COVID-19 specific subject headings became available (see Supplementary Material 1). The search strategy did not undergo formal peer review due to the need for rapid evidence early in the pandemic. The initial search was conducted from December 31, 2019 to April 13, 2020 with automated daily updates thereafter. We converted to weekly updates on December 28, 2020. The final search was done on August 31, 2023.

### Selection of Eligible Studies

Search results were imported into DistillerSR (Evidence Partners, Ottawa, Canada), where we identified and removed duplicates. Two independent reviewers evaluated titles and abstracts in random order. If either reviewer deemed a study potentially eligible, full-text review was completed by two independent reviewers with any discrepancies resolved through consensus, consulting a third reviewer as necessary. The inclusion and exclusion coding guide (Supplementary Material 2) was pre-tested, and reviewers were trained over several sessions.

### Data Extraction and Adequacy Assessment

For each eligible study, data were extracted in DistillerSR by a single reviewer using a pre-specified form with validation by a second reviewer. Reviewers extracted (1) publication characteristics (e.g., first author, year, journal); (2) population characteristics and demographics, including eligibility criteria, recruitment method, number of participants, assessment timing, age; (3) mental health outcomes including general mental health, anxiety symptoms, depression symptoms, and stress; (4) if studies reported outcomes by sex or gender or used these terms inconsistently (e.g., described using gender but reported results for females and males, which are sex terms); and (5) if sex or gender were treated as binary or categorical.

Adequacy of study methods and reporting was assessed using an adapted version of the Joanna Briggs Institute Checklist for Prevalence Studies, which assesses sampling frame appropriateness for the target population, recruiting methods, sample size, setting and participant descriptions, participation or response rate, outcome assessment methods, assessment standardization across participants, statistical analysis appropriateness, and follow-up.^9^ The 9 items were coded as “yes” for meeting adequacy criteria, “no” for not meeting criteria, or “unclear” if incomplete reporting did not allow a judgment to be made. See Supplemental Material 3.

For all data extraction, discrepancies were resolved between reviewers with a third reviewer consulted if necessary.

### Statistical Analyses

For continuous outcomes, separately for each sex or gender group, we extracted an SMD effect size with 95% CIs for change from pre-COVID-19 to during COVID-19. If not provided, we extracted pre-COVID-19 and during COVID-19 means and standard deviations (SDs) for each group, calculated raw change scores and SD, and calculated SMD for change using Hedges’ g for each group,^10^ as described by Borenstein et al.^11^ Raw change scores were presented in scale units and direction, whereas SMD change scores were presented as positive when mental health worsened from pre-COVID-19 to COVID-19 and negative when it improved. We then calculated a Hedges’ g difference in change between sex or gender groups with 95% CI. Positive numbers represented greater negative change in females or women compared to males or men.

For publications that reported proportions of participants above a scale cut-off, for pre-COVID-19 and COVID-19 proportions, if a 95% CI was not provided, we calculated a 95% CI using Agresti and Coull’s approximate method for binomial proportions.^12^ We then extracted or calculated the proportion change in participants above the cut-off, along with 95% CI, for each sex or gender group. Proportion changes were presented as positive when mental health worsened from pre-COVID-19 to COVID-19 and negative when it improved. If 95% CIs for proportion change were not reported, we generated them using Newcombe’s method for differences between binomial proportions based on paired data.^13^ This method requires the number of cases at both assessments, which was not available in some included studies. We assumed that 50% of pre-COVID-19 cases continued to be cases during COVID-19 and confirmed that results did not differ substantively if we used values from 30% to 70% (all 95% CI end points within 0.05 except in women from 1 cohort^44^; see Supplementary Table 1).^14^ Finally, we calculated a difference of the proportion change between sex or gender groups with 95% CI. Positive numbers reflected greater negative change in females or women compared to males or men.

Meta-analyses were done to synthesize differences between sex or gender groups in SMD change for continuous outcomes and proportion change for dichotomous outcomes via restricted maximum-likelihood random-effects meta-analysis. Heterogeneity was assessed with I^2^. Before the meta-analyses, multiple SMDs of studies assessing the same cohort were pooled to give the results for the cohort and that of studies with more than one measurement for a continuous outcome were pooled within the study. Meta-analyses were performed in R (R version 4.3.1, RStudio Version 2023.06.2), using the metacont and metagen functions in the meta package.^15^ Forest plots were generated using the forest function in meta.

## RESULTS

### Search Results and Selection of Eligible Studies

As of our final search on August 31, 2023, there were 149,026 unique references identified and screened for potential eligibility, of which 145,463 were excluded after title and abstract review and 2,092 after full-text review. Of 661 remaining articles with longitudinal data collection, 632 did not have eligible sex or gender data and were excluded. Thus, results from 27 unique cohorts reported in 29 publications were included. Supplementary Figure 1 shows the flow of articles and reasons for exclusion.

### Characteristics of Included Studies

Table 1 shows study characteristics. There were three adult national cohorts (five publications)^16–20^ from China (N = 18,127),^16^ the Netherlands (N = 3,983 to 4,064),^17,18^ and the United Kingdom (N = 10,918 to 15,376).^19,20^ Three cohorts ^21–23^ reported on adult community samples from Spain (N = 102),^21^ the Netherlands (N = 183),^22^ or the United States (N = 325)^23^, and one reported on an adult clinic-based sample from the United States (N = 967).^24^ Two cohorts^25,26^ reported on young adult samples; one reported on young adult twins from the United Kingdom (N = 3,563 to 3,694 depending on outcome)^25^ and another on young adults from Switzerland (N = 786).^26^ Seven cohorts^27–33^ assessed adolescents from Australia (N = 248),^27^ China (N = 831 to 1,780),^28–30^ Italy (N = 153 to 813),^31,32^ or Portugal (N = 1,099).^33^ Three cohorts^34–36^ assessed undergraduate students from China (N = 4,085 to 4,341),^34^ India (N = 217),^35^ or the United Kingdom (N = 214).^36^ Two cohorts^37,38^ assessed patients from the United States with systemic lupus erythematosus (N = 316)^37^ or moderate-to-severe traumatic brain injury (N = 365 to 694).^38^ Two cohorts^39,40^ assessed employees from petrochemical industries from China (N = 2,999)^39^ and Iran (N = 850).^40^ Five cohorts^22,23,41–43^ assessed parents from Hong Kong (N = 428),^41^ the Netherlands (N = 183),^22^ Norway (N = 309),^42^ or the United States (N = 180 to 325)^23,43^. One cohort assessed teachers from Iceland (N = 920).^44^

**Table 1.**
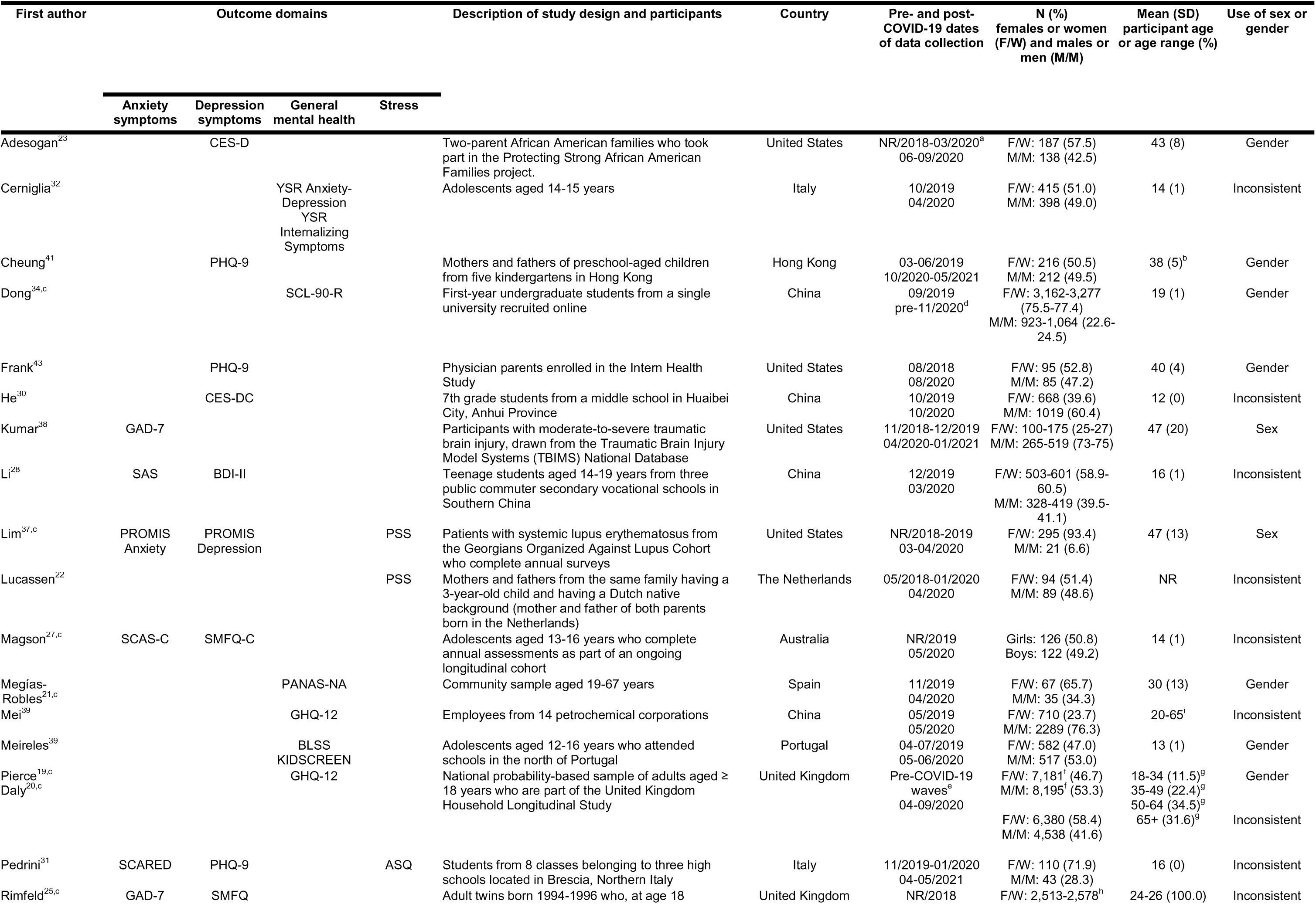
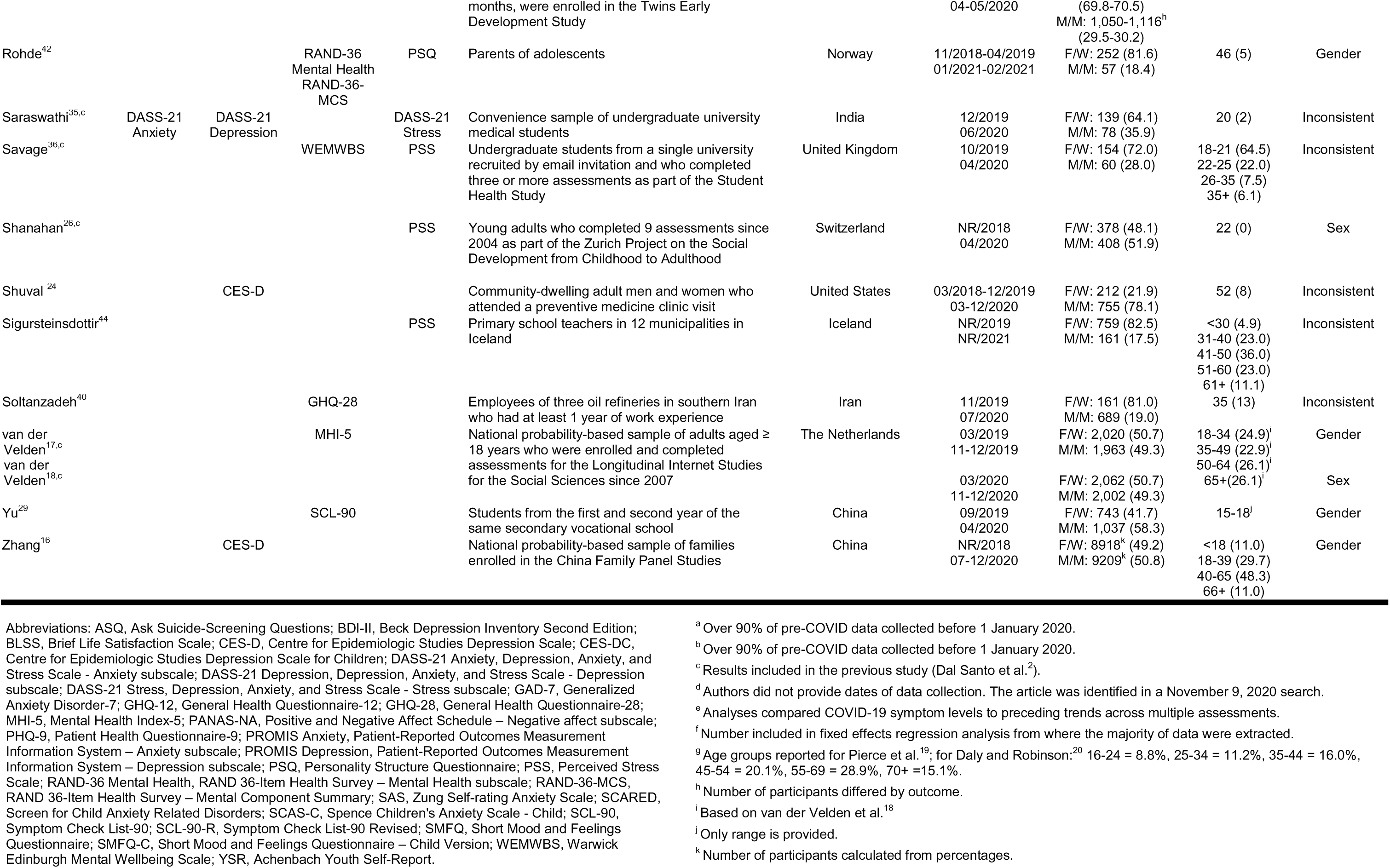
Characteristics of Included Studies (N=29).

Twelve unique cohorts (14 publications) assessed general mental health symptoms,^17–21,29,32–34,36,39–42^ 7 cohorts assessed anxiety symptoms,^25,27,28,31,35,37,38^ 12 cohorts assessed depression symptoms,^16,23–25,27,28,30,31,35,37,41,43^ and 8 cohorts assessed stress.^22,26,31,35–37,42,44^ All cohorts compared women and men or females and males; none included other sex or gender groups. Use of sex and gender terms, however, was inconsistent in 14 of 29 included publications (e.g., assessed results by gender but reported using sex terms).^20,22,24,25,27,28,30–32,35,36,39,40,44^ For assessments during COVID-19, 22 cohorts (24 publications) reported findings from 2020,^16–30,32–37,39,40,43^ and 5 cohorts reported findings from 2021.^31,38,41,42,44^

### Adequacy of Study Methods and Reporting

Among included studies, 2 publications reporting data for the same cohort were rated as “yes” for adequacy for all items.^17,18^ Other publications were rated “no” for 1–3 items (plus 0–3 unclear ratings),^16,20–24,26–36,38–44^ or “no” on none but “unclear” on 2–4 items.^19,25,37^ Overall, 22 of 29 reports were rated “no” or “unclear” for appropriate sampling frame (76%), 23 “no” or “unclear” for adequate response rate and coverage (79%), and 22 “no” or “unclear” for appropriate participant recruitment (76%). See Supplementary Table 2.

### Mental Health Symptom Changes

There were 35 total comparisons of continuous score changes and 9 of proportion changes, as shown in Table 2. Forest plots are shown in Figures 1 and 2.

**Figure 1.**
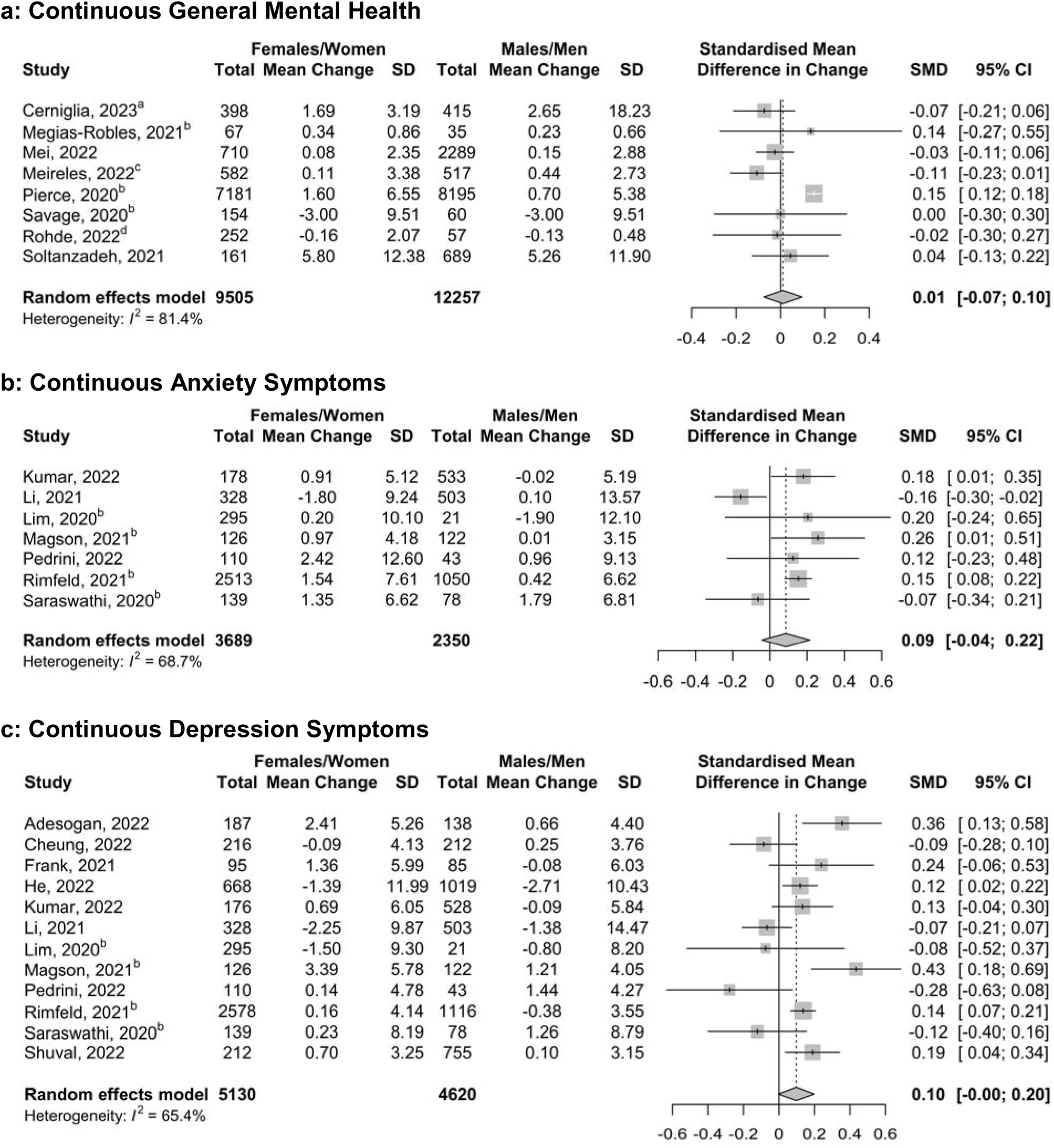
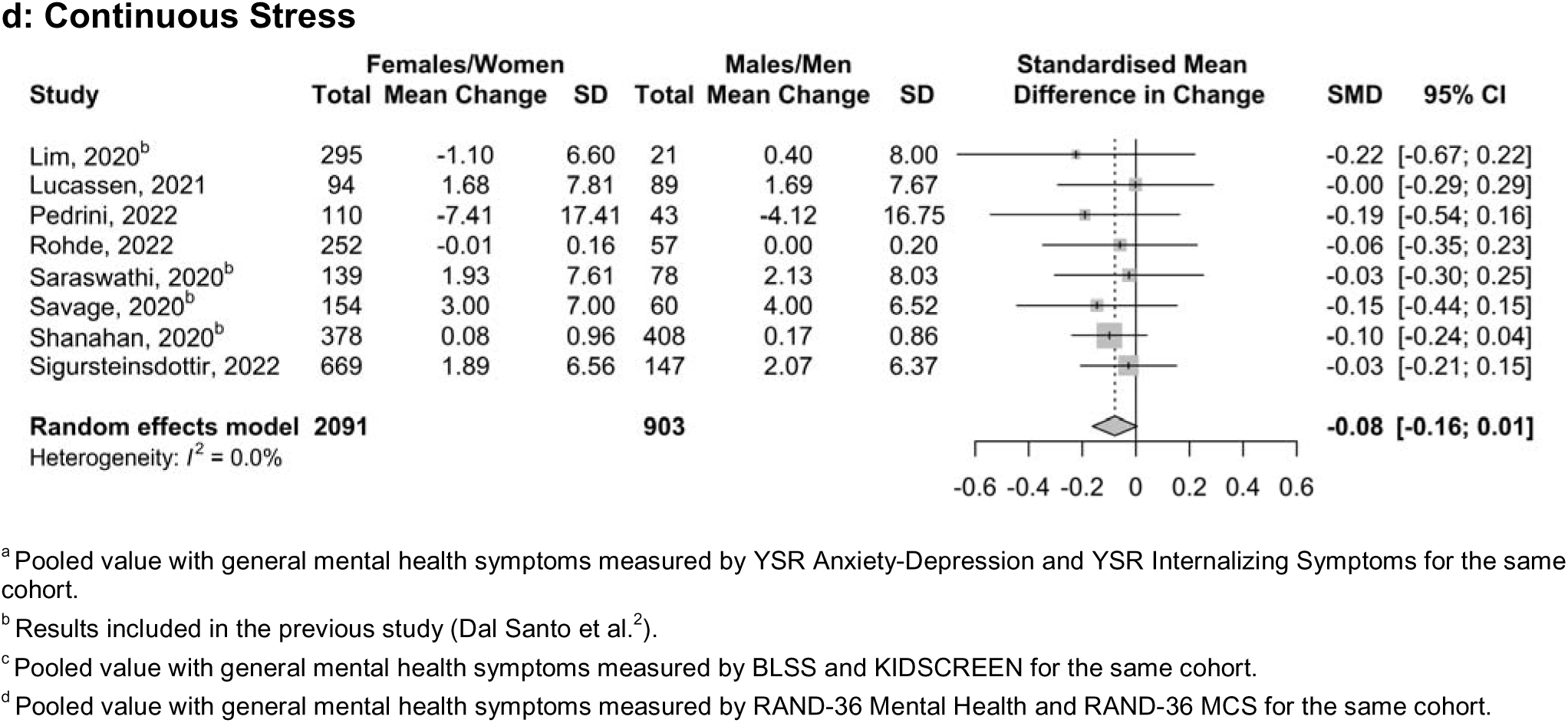
Forest Plots of Meta-Analyses for Continuous Measures of Mental Health Symptoms.

**Figure 2.**
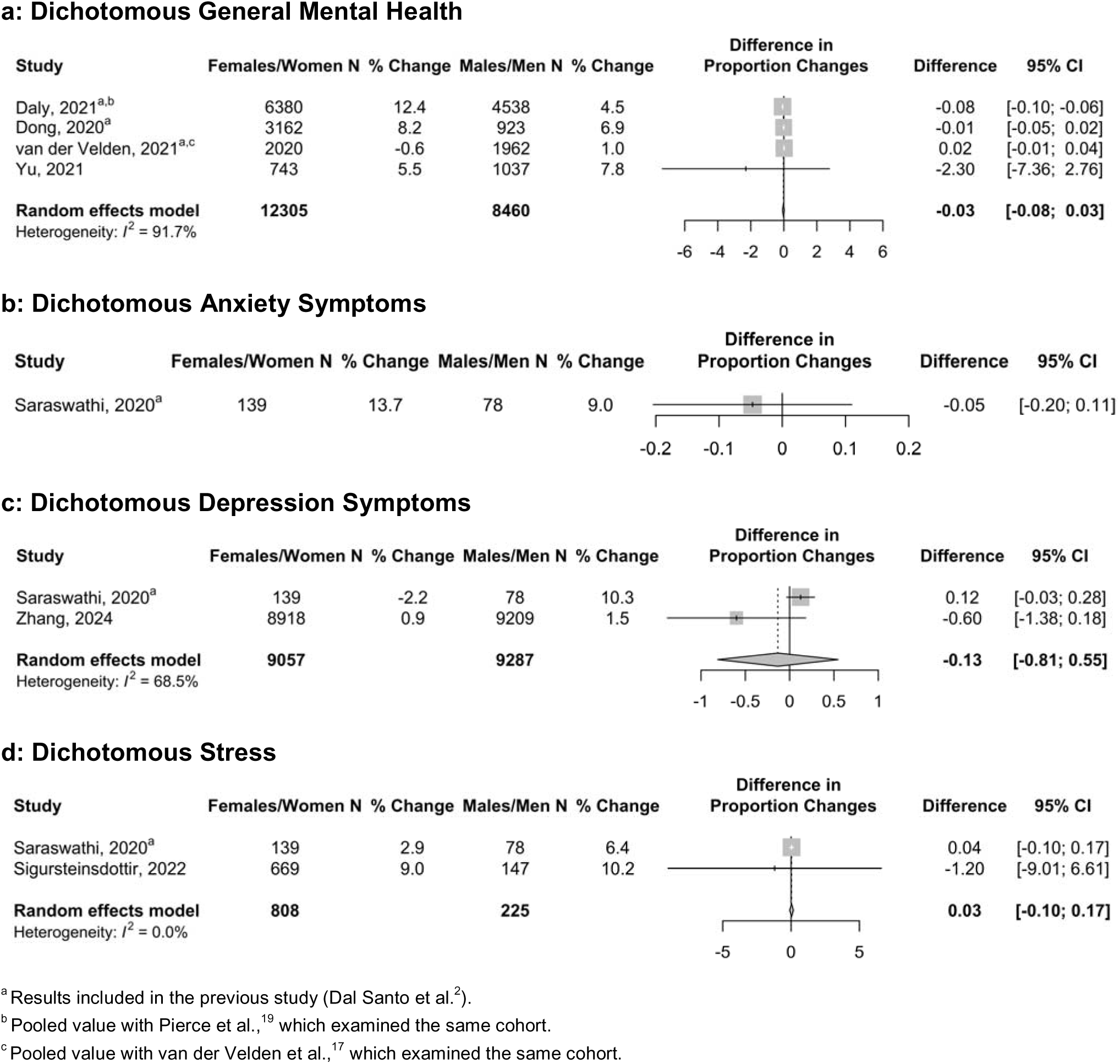
Forest Plots of Meta-Analyses for Dichotomous Measures of Mental Health Symptoms.

**Table 2.**
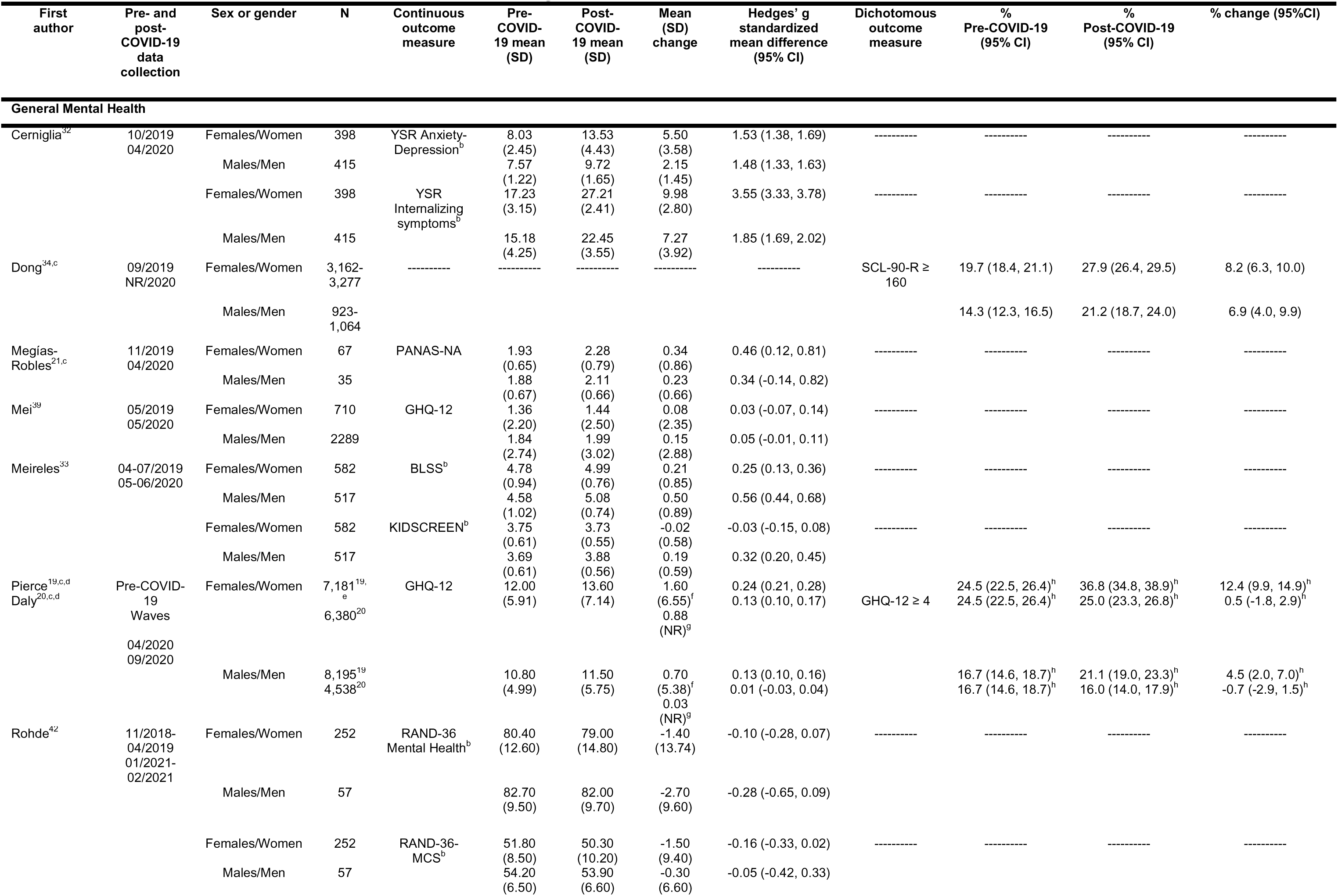
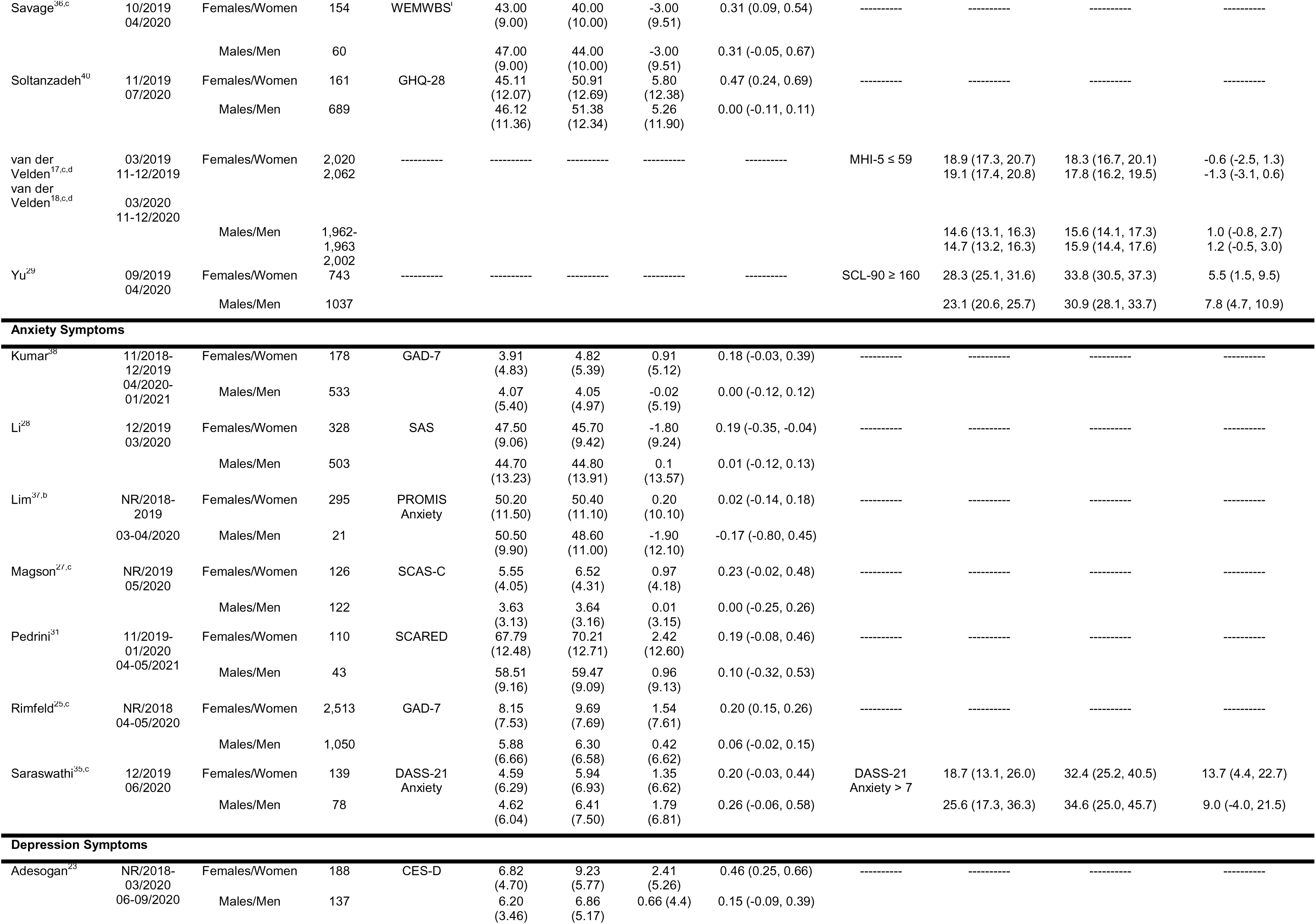
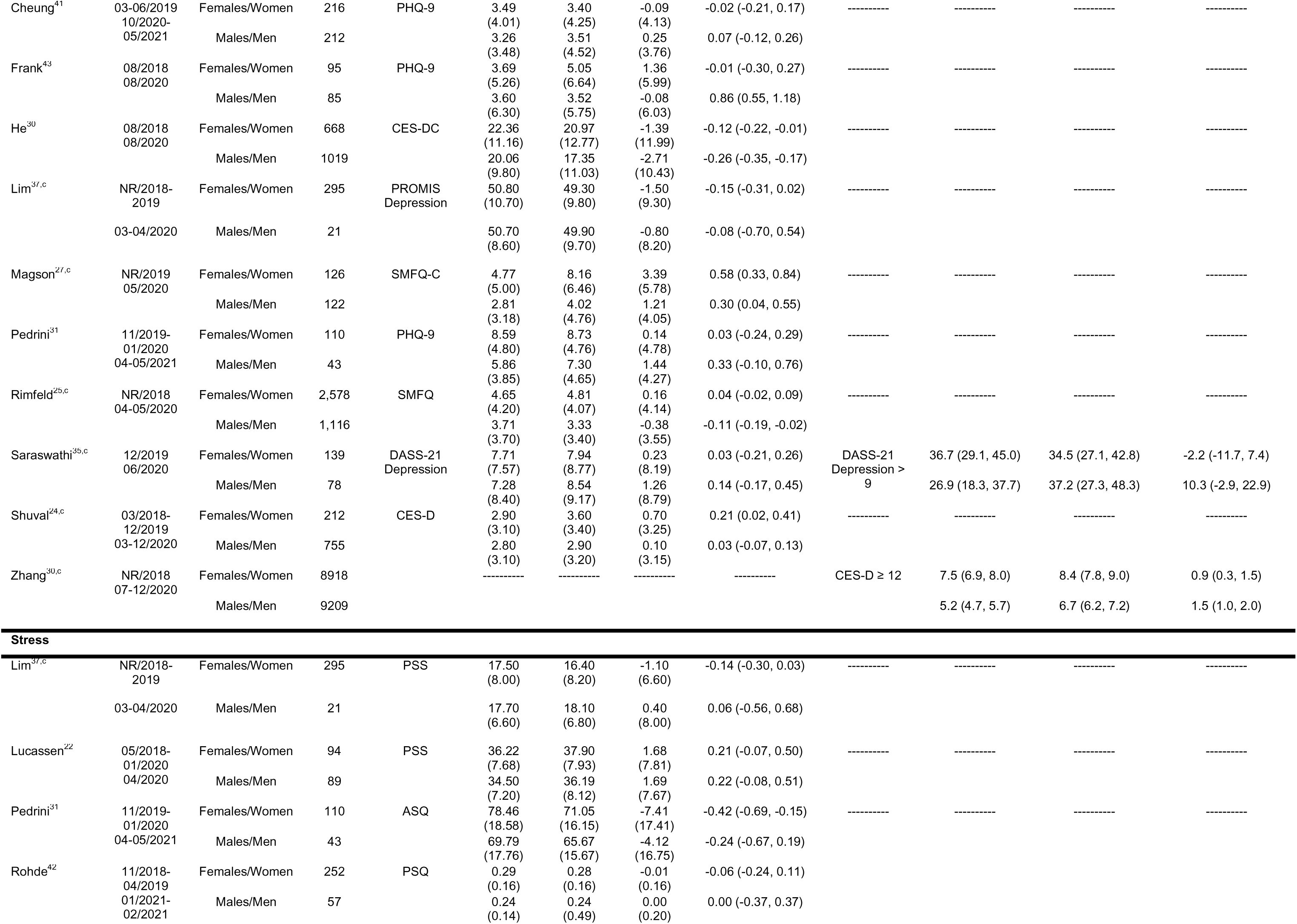
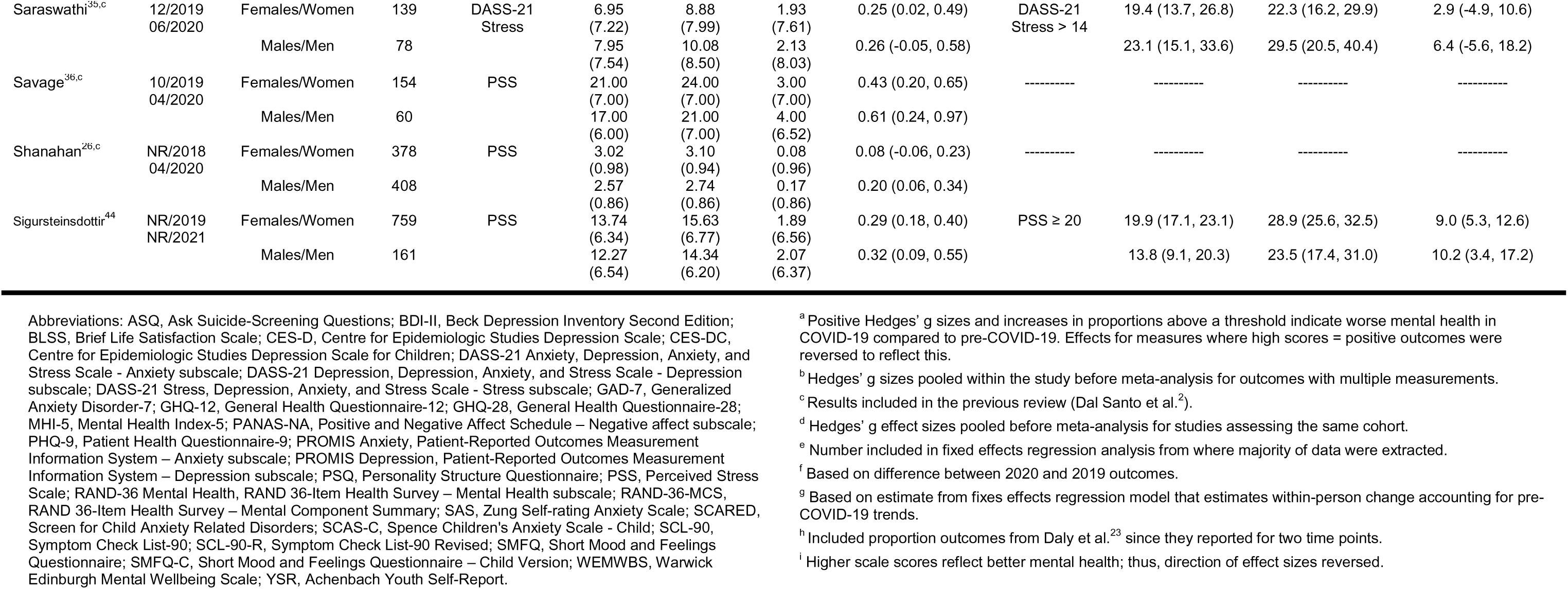
Outcomes From Included Studies by Sex or Gender^a^.

| First author | Pre- and post-COVID-19 data collection | Sex or gender | N | Continuous outcome measure | Pre-COVID-19 mean (SD) | Post-COVID-19 mean (SD) | Mean (SD) change | Hedges' g standardized mean difference (95% CI) | Dichotomous outcome measure | % Pre-COVID-19 (95% CI) | % Post-COVID-19 (95% CI) | % change (95%CI) |
| --- | --- | --- | --- | --- | --- | --- | --- | --- | --- | --- | --- | --- |
| **General Mental Health** |  |  |  |  |  |  |  |  |  |  |  |  |
| Cerniglia<sup>32</sup> | 10/2019<br>04/2020 | Females/Women | 398 | YSR Anxiety-Depression<sup>b</sup> | 8.03<br>(2.45) | 13.53<br>(4.43) | 5.50<br>(3.58) | 1.53 (1.38, 1.69) | ----- | ----- | ----- | ----- |
|  |  | Males/Men | 415 |  | 7.57<br>(1.22) | 9.72<br>(1.65) | 2.15<br>(1.45) | 1.48 (1.33, 1.63) | ----- | ----- | ----- | ----- |
|  |  | Females/Women | 398 | YSR Internalizing symptoms<sup>b</sup> | 17.23<br>(3.15) | 27.21<br>(2.41) | 9.98<br>(2.80) | 3.55 (3.33, 3.78) | ----- | ----- | ----- | ----- |
|  |  | Males/Men | 415 |  | 15.18<br>(4.25) | 22.45<br>(3.55) | 7.27<br>(3.92) | 1.85 (1.69, 2.02) | ----- | ----- | ----- | ----- |
| Dong<sup>34,c</sup> | 09/2019<br>NR/2020 | Females/Women | 3,162-3,277 | ----- | ----- | ----- | ----- | ----- | SCL-90-R ≥<br>160 | 19.7 (18.4, 21.1) | 27.9 (26.4, 29.5) | 8.2 (6.3, 10.0) |
|  |  | Males/Men | 923-1,064 | ----- | ----- | ----- | ----- | ----- |  | 14.3 (12.3, 16.5) | 21.2 (18.7, 24.0) | 6.9 (4.0, 9.9) |
| Megías-Robles<sup>21,c</sup> | 11/2019<br>04/2020 | Females/Women | 67 | PANAS-NA | 1.93<br>(0.65) | 2.28<br>(0.79) | 0.34<br>(0.86) | 0.46 (0.12, 0.81) | ----- | ----- | ----- | ----- |
|  |  | Males/Men | 35 |  | 1.88<br>(0.67) | 2.11<br>(0.66) | 0.23<br>(0.66) | 0.34 (-0.14, 0.82) | ----- | ----- | ----- | ----- |
| Mei<sup>39</sup> | 05/2019<br>05/2020 | Females/Women | 710 | GHQ-12 | 1.36<br>(2.20) | 1.44<br>(2.50) | 0.08<br>(2.35) | 0.03 (-0.07, 0.14) | ----- | ----- | ----- | ----- |
|  |  | Males/Men | 2289 |  | 1.84<br>(2.74) | 1.99<br>(3.02) | 0.15<br>(2.88) | 0.05 (-0.01, 0.11) | ----- | ----- | ----- | ----- |
| Meireles<sup>33</sup> | 04-07/2019<br>05-06/2020 | Females/Women | 582 | BLSS<sup>b</sup> | 4.78<br>(0.94) | 4.99<br>(0.76) | 0.21<br>(0.85) | 0.25 (0.13, 0.36) | ----- | ----- | ----- | ----- |
|  |  | Males/Men | 517 |  | 4.58<br>(1.02) | 5.08<br>(0.74) | 0.50<br>(0.89) | 0.56 (0.44, 0.68) | ----- | ----- | ----- | ----- |
|  |  | Females/Women | 582 | KIDSCREEN<sup>b</sup> | 3.75<br>(0.61) | 3.73<br>(0.55) | -0.02<br>(0.58) | -0.03 (-0.15, 0.08) | ----- | ----- | ----- | ----- |
|  |  | Males/Men | 517 |  | 3.69<br>(0.61) | 3.88<br>(0.56) | 0.19<br>(0.59) | 0.32 (0.20, 0.45) | ----- | ----- | ----- | ----- |
| Pierce<sup>19,c,d</sup><br>Daly<sup>20,c,d</sup> | Pre-COVID-19<br>Waves<br>04/2020<br>09/2020 | Females/Women | 7,181<sup>19</sup><br><sub>e</sub><br>6,380<sup>20</sup> | GHQ-12 | 12.00<br>(5.91) | 13.60<br>(7.14) | 1.60<br>(6.55)<sup>f</sup><br>0.88<br>(NR)<sup>g</sup> | 0.24 (0.21, 0.28)<br>0.13 (0.10, 0.17) | GHQ-12 ≥ 4 | 24.5 (22.5, 26.4)<sup>h</sup><br>24.5 (22.5, 26.4)<sup>h</sup> | 36.8 (34.8, 38.9)<sup>h</sup><br>25.0 (23.3, 26.8)<sup>h</sup> | 12.4 (9.9, 14.9)<sup>h</sup><br>0.5 (-1.8, 2.9)<sup>h</sup> |
|  |  | Males/Men | 8,195<sup>19</sup><br>4,538<sup>20</sup> |  | 10.80<br>(4.99) | 11.50<br>(5.75) | 0.70<br>(5.38)<sup>f</sup><br>0.03<br>(NR)<sup>g</sup> | 0.13 (0.10, 0.16)<br>0.01 (-0.03, 0.04) |  | 16.7 (14.6, 18.7)<sup>h</sup><br>16.7 (14.6, 18.7)<sup>h</sup> | 21.1 (19.0, 23.3)<sup>h</sup><br>16.0 (14.0, 17.9)<sup>h</sup> | 4.5 (2.0, 7.0)<sup>h</sup><br>-0.7 (-2.9, 1.5)<sup>h</sup> |
| Rohde<sup>42</sup> | 11/2018-04/2019<br>01/2021-02/2021 | Females/Women | 252 | RAND-36<br>Mental Health<sup>b</sup> | 80.40<br>(12.60) | 79.00<br>(14.80) | -1.40<br>(13.74) | -0.10 (-0.28, 0.07) | ----- | ----- | ----- | ----- |
|  |  | Males/Men | 57 |  | 82.70<br>(9.50) | 82.00<br>(9.70) | -2.70<br>(9.60) | -0.28 (-0.65, 0.09) | ----- | ----- | ----- | ----- |
|  |  | Females/Women | 252 | RAND-36-MCS<sup>b</sup> | 51.80<br>(8.50) | 50.30<br>(10.20) | -1.50<br>(9.40) | -0.16 (-0.33, 0.02) | ----- | ----- | ----- | ----- |
|  |  | Males/Men | 57 |  | 54.20<br>(6.50) | 53.90<br>(6.60) | -0.30<br>(6.60) | -0.05 (-0.42, 0.33) | ----- | ----- | ----- | ----- |
| Savage<sup>36,c</sup> | 10/2019<br>04/2020 | Females/Women | 154 | WEMWBS<sup>i</sup> | 43.00<br>(9.00) | 40.00<br>(10.00) | -3.00<br>(9.51) | 0.31 (0.09, 0.54) | ----- | ----- | ----- | ----- |
|  |  | Males/Men | 60 |  | 47.00<br>(9.00) | 44.00<br>(10.00) | -3.00<br>(9.51) | 0.31 (-0.05, 0.67) |  |  |  |  |
| Soltanzadeh<sup>40</sup> | 11/2019<br>07/2020 | Females/Women | 161 | GHQ-28 | 45.11<br>(12.07) | 50.91<br>(12.69) | 5.80<br>(12.38) | 0.47 (0.24, 0.69) | ----- | ----- | ----- | ----- |
|  |  | Males/Men | 689 |  | 46.12<br>(11.36) | 51.38<br>(12.34) | 5.26<br>(11.90) | 0.00 (-0.11, 0.11) |  |  |  |  |
| van der Velden<sup>17,c,d</sup><br>van der Velden<sup>18,c,d</sup> | 03/2019<br>11-12/2019 | Females/Women | 2,020<br>2,062 | ----- | ----- | ----- | ----- | ----- | MHI-5 ≤ 59 | 18.9 (17.3, 20.7)<br>19.1 (17.4, 20.8) | 18.3 (16.7, 20.1)<br>17.8 (16.2, 19.5) | -0.6 (-2.5, 1.3)<br>-1.3 (-3.1, 0.6) |
|  | 03/2020<br>11-12/2020 | Males/Men | 1,962-1,963<br>2,002 |  |  |  |  |  |  | 14.6 (13.1, 16.3)<br>14.7 (13.2, 16.3) | 15.6 (14.1, 17.3)<br>15.9 (14.4, 17.6) | 1.0 (-0.8, 2.7)<br>1.2 (-0.5, 3.0) |
| Yu<sup>29</sup> | 09/2019<br>04/2020 | Females/Women | 743 | ----- | ----- | ----- | ----- | ----- | SCL-90 ≥ 160 | 28.3 (25.1, 31.6) | 33.8 (30.5, 37.3) | 5.5 (1.5, 9.5) |
|  |  | Males/Men | 1037 |  |  |  |  |  |  | 23.1 (20.6, 25.7) | 30.9 (28.1, 33.7) | 7.8 (4.7, 10.9) |
| **Anxiety Symptoms** |  |  |  |  |  |  |  |  |  |  |  |  |
| Kumar<sup>38</sup> | 11/2018-12/2019 | Females/Women | 178 | GAD-7 | 3.91<br>(4.83) | 4.82<br>(5.39) | 0.91<br>(5.12) | 0.18 (-0.03, 0.39) | ----- | ----- | ----- | ----- |
|  | 04/2020-01/2021 | Males/Men | 533 |  | 4.07<br>(5.40) | 4.05<br>(4.97) | -0.02<br>(5.19) | 0.00 (-0.12, 0.12) |  |  |  |  |
| Li<sup>28</sup> | 12/2019<br>03/2020 | Females/Women | 328 | SAS | 47.50<br>(9.06) | 45.70<br>(9.42) | -1.80<br>(9.24) | 0.19 (-0.35, -0.04) | ----- | ----- | ----- | ----- |
|  |  | Males/Men | 503 |  | 44.70<br>(13.23) | 44.80<br>(13.91) | 0.1<br>(13.57) | 0.01 (-0.12, 0.13) |  |  |  |  |
| Lim<sup>37,b</sup> | NR/2018-2019 | Females/Women | 295 | PROMIS<br>Anxiety | 50.20<br>(11.50) | 50.40<br>(11.10) | 0.20<br>(10.10) | 0.02 (-0.14, 0.18) | ----- | ----- | ----- | ----- |
|  | 03-04/2020 | Males/Men | 21 |  | 50.50<br>(9.90) | 48.60<br>(11.00) | -1.90<br>(12.10) | -0.17 (-0.80, 0.45) |  |  |  |  |
| Magson<sup>27,c</sup> | NR/2019<br>05/2020 | Females/Women | 126 | SCAS-C | 5.55<br>(4.05) | 6.52<br>(4.31) | 0.97<br>(4.18) | 0.23 (-0.02, 0.48) | ----- | ----- | ----- | ----- |
|  |  | Males/Men | 122 |  | 3.63<br>(3.13) | 3.64<br>(3.16) | 0.01<br>(3.15) | 0.00 (-0.25, 0.26) |  |  |  |  |
| Pedrini<sup>31</sup> | 11/2019-01/2020 | Females/Women | 110 | SCARED | 67.79<br>(12.48) | 70.21<br>(12.71) | 2.42<br>(12.60) | 0.19 (-0.08, 0.46) | ----- | ----- | ----- | ----- |
|  | 04-05/2021 | Males/Men | 43 |  | 58.51<br>(9.16) | 59.47<br>(9.09) | 0.96<br>(9.13) | 0.10 (-0.32, 0.53) |  |  |  |  |
| Rimfeld<sup>25,c</sup> | NR/2018<br>04-05/2020 | Females/Women | 2,513 | GAD-7 | 8.15<br>(7.53) | 9.69<br>(7.69) | 1.54<br>(7.61) | 0.20 (0.15, 0.26) | ----- | ----- | ----- | ----- |
|  |  | Males/Men | 1,050 |  | 5.88<br>(6.66) | 6.30<br>(6.58) | 0.42<br>(6.62) | 0.06 (-0.02, 0.15) |  |  |  |  |
| Saraswathi<sup>35,c</sup> | 12/2019<br>06/2020 | Females/Women | 139 | DASS-21<br>Anxiety | 4.59<br>(6.29) | 5.94<br>(6.93) | 1.35<br>(6.62) | 0.20 (-0.03, 0.44) | DASS-21<br>Anxiety > 7 | 18.7 (13.1, 26.0) | 32.4 (25.2, 40.5) | 13.7 (4.4, 22.7) |
|  |  | Males/Men | 78 |  | 4.62<br>(6.04) | 6.41<br>(7.50) | 1.79<br>(6.81) | 0.26 (-0.06, 0.58) |  | 25.6 (17.3, 36.3) | 34.6 (25.0, 45.7) | 9.0 (-4.0, 21.5) |
| **Depression Symptoms** |  |  |  |  |  |  |  |  |  |  |  |  |
| Adesogan<sup>23</sup> | NR/2018-03/2020 | Females/Women | 188 | CES-D | 6.82<br>(4.70) | 9.23<br>(5.77) | 2.41<br>(5.26) | 0.46 (0.25, 0.66) | ----- | ----- | ----- | ----- |
|  | 06-09/2020 | Males/Men | 137 |  | 6.20<br>(3.46) | 6.86<br>(5.17) | 0.66 (4.4) | 0.15 (-0.09, 0.39) |  |  |  |  |
| Cheung<sup>41</sup> | 03-06/2019<br>10/2020-05/2021 | Females/Women | 216 | PHQ-9 | 3.49<br>(4.01) | 3.40<br>(4.25) | -0.09<br>(4.13) | -0.02 (-0.21, 0.17) | ----- | ----- | ----- | ----- |
|  |  | Males/Men | 212 |  | 3.26<br>(3.48) | 3.51<br>(4.52) | 0.25<br>(3.76) | 0.07 (-0.12, 0.26) | ----- | ----- | ----- | ----- |
| Frank<sup>43</sup> | 08/2018<br>08/2020 | Females/Women | 95 | PHQ-9 | 3.69<br>(5.26) | 5.05<br>(6.64) | 1.36<br>(5.99) | -0.01 (-0.30, 0.27) | ----- | ----- | ----- | ----- |
|  |  | Males/Men | 85 |  | 3.60<br>(6.30) | 3.52<br>(5.75) | -0.08<br>(6.03) | 0.86 (0.55, 1.18) | ----- | ----- | ----- | ----- |
| He<sup>30</sup> | 08/2018<br>08/2020 | Females/Women | 668 | CES-DC | 22.36<br>(11.16) | 20.97<br>(12.77) | -1.39<br>(11.99) | -0.12 (-0.22, -0.01) | ----- | ----- | ----- | ----- |
|  |  | Males/Men | 1019 |  | 20.06<br>(9.80) | 17.35<br>(11.03) | -2.71<br>(10.43) | -0.26 (-0.35, -0.17) | ----- | ----- | ----- | ----- |
| Lim<sup>37,c</sup> | NR/2018-2019 | Females/Women | 295 | PROMIS<br>Depression | 50.80<br>(10.70) | 49.30<br>(9.80) | -1.50<br>(9.30) | -0.15 (-0.31, 0.02) | ----- | ----- | ----- | ----- |
|  |  | Males/Men | 21 |  | 50.70<br>(8.60) | 49.90<br>(9.70) | -0.80<br>(8.20) | -0.08 (-0.70, 0.54) | ----- | ----- | ----- | ----- |
| Magson<sup>27,c</sup> | NR/2019<br>05/2020 | Females/Women | 126 | SMFQ-C | 4.77<br>(5.00) | 8.16<br>(6.46) | 3.39<br>(5.78) | 0.58 (0.33, 0.84) | ----- | ----- | ----- | ----- |
|  |  | Males/Men | 122 |  | 2.81<br>(3.18) | 4.02<br>(4.76) | 1.21<br>(4.05) | 0.30 (0.04, 0.55) | ----- | ----- | ----- | ----- |
| Pedrini<sup>31</sup> | 11/2019-01/2020<br>04-05/2021 | Females/Women | 110 | PHQ-9 | 8.59<br>(4.80) | 8.73<br>(4.76) | 0.14<br>(4.78) | 0.03 (-0.24, 0.29) | ----- | ----- | ----- | ----- |
|  |  | Males/Men | 43 |  | 5.86<br>(3.85) | 7.30<br>(4.65) | 1.44<br>(4.27) | 0.33 (-0.10, 0.76) | ----- | ----- | ----- | ----- |
| Rimfeld<sup>25,c</sup> | NR/2018<br>04-05/2020 | Females/Women | 2,578 | SMFQ | 4.65<br>(4.20) | 4.81<br>(4.07) | 0.16<br>(4.14) | 0.04 (-0.02, 0.09) | ----- | ----- | ----- | ----- |
|  |  | Males/Men | 1,116 |  | 3.71<br>(3.70) | 3.33<br>(3.40) | -0.38<br>(3.55) | -0.11 (-0.19, -0.02) | ----- | ----- | ----- | ----- |
| Saraswathi<sup>35,c</sup> | 12/2019<br>06/2020 | Females/Women | 139 | DASS-21<br>Depression | 7.71<br>(7.57) | 7.94<br>(8.77) | 0.23<br>(8.19) | 0.03 (-0.21, 0.26) | DASS-21<br>Depression ><br>9 | 36.7 (29.1, 45.0) | 34.5 (27.1, 42.8) | -2.2 (-11.7, 7.4) |
|  |  | Males/Men | 78 |  | 7.28<br>(8.40) | 8.54<br>(9.17) | 1.26<br>(8.79) | 0.14 (-0.17, 0.45) |  | 26.9 (18.3, 37.7) | 37.2 (27.3, 48.3) | 10.3 (-2.9, 22.9) |
| Shuval<sup>24,c</sup> | 03/2018-12/2019<br>03-12/2020 | Females/Women | 212 | CES-D | 2.90<br>(3.10) | 3.60<br>(3.40) | 0.70<br>(3.25) | 0.21 (0.02, 0.41) | ----- | ----- | ----- | ----- |
|  |  | Males/Men | 755 |  | 2.80<br>(3.10) | 2.90<br>(3.20) | 0.10<br>(3.15) | 0.03 (-0.07, 0.13) | ----- | ----- | ----- | ----- |
| Zhang<sup>30,c</sup> | NR/2018<br>07-12/2020 | Females/Women | 8918 |  | ----- | ----- | ----- | ----- | CES-D ≥ 12 | 7.5 (6.9, 8.0) | 8.4 (7.8, 9.0) | 0.9 (0.3, 1.5) |
|  |  | Males/Men | 9209 |  | ----- | ----- | ----- | ----- |  | 5.2 (4.7, 5.7) | 6.7 (6.2, 7.2) | 1.5 (1.0, 2.0) |
| Stress |  |  |  |  |  |  |  |  |  |  |  |  |
| Lim<sup>37,c</sup> | NR/2018-2019 | Females/Women | 295 | PSS | 17.50<br>(8.00) | 16.40<br>(8.20) | -1.10<br>(6.60) | -0.14 (-0.30, 0.03) | ----- | ----- | ----- | ----- |
|  |  | Males/Men | 21 |  | 17.70<br>(6.60) | 18.10<br>(6.80) | 0.40<br>(8.00) | 0.06 (-0.56, 0.68) | ----- | ----- | ----- | ----- |
| Lucassen<sup>22</sup> | 05/2018-01/2020<br>04/2020 | Females/Women | 94 | PSS | 36.22<br>(7.68) | 37.90<br>(7.93) | 1.68<br>(7.81) | 0.21 (-0.07, 0.50) | ----- | ----- | ----- | ----- |
|  |  | Males/Men | 89 |  | 34.50<br>(7.20) | 36.19<br>(8.12) | 1.69<br>(7.67) | 0.22 (-0.08, 0.51) | ----- | ----- | ----- | ----- |
| Pedrini<sup>31</sup> | 11/2019-01/2020<br>04-05/2021 | Females/Women | 110 | ASQ | 78.46<br>(18.58) | 71.05<br>(16.15) | -7.41<br>(17.41) | -0.42 (-0.69, -0.15) | ----- | ----- | ----- | ----- |
|  |  | Males/Men | 43 |  | 69.79<br>(17.76) | 65.67<br>(15.67) | -4.12<br>(16.75) | -0.24 (-0.67, 0.19) | ----- | ----- | ----- | ----- |
| Rohde<sup>42</sup> | 11/2018-04/2019<br>01/2021-02/2021 | Females/Women | 252 | PSQ | 0.29<br>(0.16) | 0.28<br>(0.16) | -0.01<br>(0.16) | -0.06 (-0.24, 0.11) | ----- | ----- | ----- | ----- |
|  |  | Males/Men | 57 |  | 0.24<br>(0.14) | 0.24<br>(0.49) | 0.00<br>(0.20) | 0.00 (-0.37, 0.37) | ----- | ----- | ----- | ----- |
| Saraswathi<sup>35,c</sup> | 12/2019<br>06/2020 | Females/Women | 139 | DASS-21<br>Stress | 6.95<br>(7.22) | 8.88<br>(7.99) | 1.93<br>(7.61) | 0.25 (0.02, 0.49) | DASS-21<br>Stress > 14 | 19.4 (13.7, 26.8) | 22.3 (16.2, 29.9) | 2.9 (-4.9, 10.6) |
|  |  | Males/Men | 78 |  | 7.95<br>(7.54) | 10.08<br>(8.50) | 2.13<br>(8.03) | 0.26 (-0.05, 0.58) |  | 23.1 (15.1, 33.6) | 29.5 (20.5, 40.4) | 6.4 (-5.6, 18.2) |
| Savage<sup>36,c</sup> | 10/2019<br>04/2020 | Females/Women | 154 | PSS | 21.00<br>(7.00) | 24.00<br>(7.00) | 3.00<br>(7.00) | 0.43 (0.20, 0.65) | ----- | ----- | ----- | ----- |
|  |  | Males/Men | 60 |  | 17.00<br>(6.00) | 21.00<br>(7.00) | 4.00<br>(6.52) | 0.61 (0.24, 0.97) |  |  |  |  |
| Shanahan<sup>26,c</sup> | NR/2018<br>04/2020 | Females/Women | 378 | PSS | 3.02<br>(0.98) | 3.10<br>(0.94) | 0.08<br>(0.96) | 0.08 (-0.06, 0.23) | ----- | ----- | ----- | ----- |
|  |  | Males/Men | 408 |  | 2.57<br>(0.86) | 2.74<br>(0.86) | 0.17<br>(0.86) | 0.20 (0.06, 0.34) |  |  |  |  |
| Sigursteinsdottir<sup>44</sup> | NR/2019<br>NR/2021 | Females/Women | 759 | PSS | 13.74<br>(6.34) | 15.63<br>(6.77) | 1.89<br>(6.56) | 0.29 (0.18, 0.40) | PSS ≥ 20 | 19.9 (17.1, 23.1) | 28.9 (25.6, 32.5) | 9.0 (5.3, 12.6) |
|  |  | Males/Men | 161 |  | 12.27<br>(6.54) | 14.34<br>(6.20) | 2.07<br>(6.37) | 0.32 (0.09, 0.55) |  | 13.8 (9.1, 20.3) | 23.5 (17.4, 31.0) | 10.2 (3.4, 17.2) |

Estimates of differences in change by sex or gender were close to zero and not statistically significant for continuously measured outcomes, including general mental health (Figure 1a; SMD change_women-men_□=□0.01, 95% CI□0.07 to 0.10; N□= 8 cohorts,^19,21,32,33,36,39,40,42^ 21,762 participants; I^2^□=□81%), anxiety symptoms (Figure 1b; SMD change_women-men_□=□0.09, 95% CI□0.04 to 0.22; N□= 7 cohorts,^25,27,28,31,35,37,38^ 6,039 participants; I^2^□=□69%), depression symptoms (Figure 1c; SMD change_women-men_□= 0.10, 95% CI −0.00 to 0.20; N□= 12 cohorts,^23–25,27,28,30,31,35,37,38,41,43^ 9,750 participants; I^2^□=□65%), and stress (Figure 1d; SMD change_women-men_□=□-0.08, 95% CI□0.16 to 0.01; N□= 8 cohorts,^22,26,31,35–37,42,44^ 2,994 participants; I^2^□=□0%).

No dichotomous changes were statistically significant: general mental health (Figure 2a; proportion change_women-men_ □= −0.03, 95% CI −0.08 to 0.03; N□= 4 cohorts,^17,20,29,34^ 20,765 participants, I^2^ = 92%), anxiety symptoms (Figure 2b; proportion change_women-men_ □= −0.05, 95% CI −0.20 to 0.11; N□= 1 cohort,^35^ 217 participants), depression symptoms (Figure 2c; proportion change_women-men_ □= −0.13, 95% CI −0.81 to 0.55; N□=□2 cohorts,^16,35^ 18,344 participants, I^2^ = 69%), and stress (Figure 2d; proportion change_women-men_ □= 0.04, 95% CI −0.10 to 0.17; N□= 2 cohorts,^35,44^ 1,033 participants, I^2^ = 0%).

## DISCUSSION

We reviewed evidence from 27 unique cohorts that reported longitudinal changes in mental health symptoms from before to during the COVID-19 pandemic by sex or gender. We found comparisons of females or women versus males or men but no other sex or gender groups. We did not find any substantive or statistically significant differences in changes by sex or gender, which differs from our initial meta-analysis of 12 cohorts that found that females or women worsened more than males or men in continuously measured anxiety and general mental health symptoms but not in depression symptoms or stress.^2^

### Comparison with Other Studies

Earlier in the pandemic, women or females were anticipated to experience greater worsening in mental health as compared to men or males due to factors including pregnancy status, domestic violence, heavier household and childcare burdens, and worsened socioeconomic environment.^45–50^ To our knowledge, three systematic reviews and meta-analyses compared data before and during the pandemic, two reported significant differences in mental health changes across sex or gender for depression and anxiety disorders as well as psychological distress,^51,52^ while one found no gender differences in suicide rates.^53^ Both reviews with significant gendered differences relied mostly on cross-sectional studies but very few longitudinal ones. The first review analyzed 48 population surveys reporting prevalences of major depressive and anxiety disorders and estimated change in prevalence by logistic meta-regression. They found females to be more affected than males for both depression (% increase in prevalence in females = 29.8, 95% CI [27.3–32.5], vs. in males = 24.0, 95% CI [21.5–26.7]) and anxiety (% increase in prevalence in females = 27.9, 95% CI [25.6–30.4], vs. in males = 21.7, 95% CI [19.3–24.1]).^51^ The other meta-analyzed 21 studies estimating the prevalence of psychological distress due to the COVID-19 pandemic with the Psychological Distress Impact tool and concluded that the overall pooled prevalence of distress due to the pandemic were significantly higher in females (53.0%) than in males (39.0%).^52^ Another systematic review included 26 studies reported suicide statistics by sex, where most studies find none or minimal difference between sexes. Meta-analyses were not conducted possibly due to significant heterogeneity in suicide data reporting of included studies.^53^

The difference between our findings and previous studies may be attributable to the use of cross-sectional rather than longitudinal data in those studies.^1^ When measured cross-sectionally, women or females consistently report substantially higher levels of mental health symptoms than males or men.^54,55^ Cross-sectional studies capture these differences, but they cannot disentangle existing sex or gender differences from COVID-19-specific changes. To isolate pandemic-related changes, we included only longitudinal studies with pre-COVID-19 and during-COVID-19 assessments and, thus, we excluded most of the studies evaluated in the two prior reviews of mental health symptoms that reported significant gendered effects.^51,52^

### Strengths and Limitations

Strengths of our study include the inclusion of data from longitudinal comparisons of change instead of cross-sectional comparisons of levels, which avoids conflation of ongoing differences with change from pre-pandemic’s to during the pandemic. Additionally, we searched 10 databases, including Chinese-language databases, without language restrictions. We also implemented well-defined inclusion criteria, such as specific thresholds for proportions of population assessed and management of losses to follow-up, to support interpretability of change estimates.

There are also limitations to consider. First, the geographic representativeness of our study is limited. Of 27 included cohorts, 18 were from high-income countries, 7 from China, and 2 from other Asian countries. We did not identify eligible studies from African, South American, and other countries. Second, we were unable to conduct subgroup analyses based on additional sociodemographic or other factors since only 1 of 27 cohorts reported mental health symptom changes by ethnicity or race^38^ and 2 by age group,^40,44^ for instance. Third, none of the included cohorts reported results from gender-diverse individuals or gender minority groups.

Sex and gender are well-recognized important determinants of health and wellbeing, and although gender minority groups may have been at risk for psychological distress and worsening of mental health symptoms during the pandemic,^56,57^ none of the cohorts we were able to include recorded outcomes for gender-diverse individuals as suggested by the Sex and Gender Equity in Research (SAGER) guidelines.^58^

## CONCLUSION

We did not find evidence that the COVID-19 pandemic’s impact on mental health differed across sex or gender groups. Our findings contrast with some cross-sectional studies, highlighting the importance of longitudinal analysis in assessing pandemic-related mental health changes. The lack of data from gender-diverse groups emphasizes the need for adherence to SAGER guidelines to ensure reporting of data from sex and gender minority groups, even if numbers are small, to facilitate synthesis. Future research should address the underrepresentation of developing countries and include data on gender-diverse individuals to provide a more comprehensive understanding of mental health symptom changes across diverse populations.

## Ethical Approval

Research ethics approval was not required since the study involved previously published summary statistics.

## Authors’ contributions

**Conception and design:** YWu, YS, AB, BDT; **data acquisition:** GCYS, YWu, SF, TDS, NPG, LL, KL, XJ, AT, YWang, JTB, PD, BT, MDO, AK, CA, CH, RSH, AA, OB, DBR, SM, MA, EN, SH, MC, DN, LA, IT, AB, BDT; **data analysis:** GCYS, YWu, SF, TDS, NPG, BDT; **article writing:** GCYS, YWu, SF, TDS, BDT; **final approval:** YWu, AB, BDT.

## Data Availability

All data produced in the present work are contained in the manuscript.

