## Supplementa Materials for "Mental health symptom changes by sex or gender during COVID-19 compared to pre-pandemic: a systematic review and meta-analysis update"

**SUPPLEMENTAL MATERIAL**

**Table of Contents**

**Supplemental Material 1**. Search strategy

**Supplemental Material 2.** Title and Abstract and Full-Text Review Inclusion and Exclusion Criteria Coding Guides

**Supplemental Material 3.** Adequacy of Study Methods and Reporting – Adapted Joanna Briggs Institute Checklist for Prevalence Studies

**Supplementary Table 1.** Confidence Intervals for Proportion Change When Assuming that 30% and 70% of Pre-COVID-19 Cases were Cases During COVID-19

**Supplementary Figure 1.** PRISMA Flowchart

**Supplementary Table 2.** Adequacy of Methods and Reporting of Included Studies (N=29)

**Supplemental Material 1**. Search strategy

Ovid MEDLINE All

†New subject heading added to original search on January 27, 2021

1. Quarantine/

2. social isolation/ or loneliness/ or physical distancing/†

3. psychology.fs. or psychology/

4. Mental Health/

5. mental disorders/

6. social stigma/

7. Fear/

8. Anxiety/

9. Depression/

10. Stress, Physiological/ or Stress, Psychological/

11. Anger/

12. Irritable Mood/

13. Grief/

14. burnout, psychological/ or burnout, professional/

15. or/1-14

16. (Quarantine* or Self-isolation or isolation or social distanc* or shelter*-in-place or psych* or mental health or mental illness* or mental disorder* or stigma or fear* or anxiety or anxious or depression or depressive or loneliness or stress* or trauma* or post-traumatic or posttraumatic or anger or mood* or irritability or irritable or emotional disturbance* or grief or burned out or burnout).tw,kf.

17. ((exp coronavirus/ or exp coronavirus infections/ or (betacoronavirus* or beta coronavirus* or coronavirus* or corona virus*).mp.) and (exp china/ or (china or chinese or hubei or wuhan).af.)) or (coronavirus* or corona virus* or betacoronavirus* or beta coronavirus*).mp.

18. (severe acute respiratory syndrome coronavirus 2 or "SARS CoV-2" or "SARSCoV 2" or SARSCoV2 or cov2 or "sars 2" or COVID or "coronavirus 2" or covid19 or nCov or ((new or Novel) adj3 coronavirus*) or ncp).mp. or ((exp pneumonia/ or pneumonia.mp.) and wuhan.af.)

19. 17 or 18

20. 15 or 16

21. 19 and 20

22. ("20191231" or 2020* or 2021*).dt,ez,da.

23. 21 and 22

Embase (Ovid)

1. exp coronavirinae/

2. exp Coronavirus infection/

3. (betacoronavirus* or beta coronavirus* or coronavirus* or corona virus*).mp.

4. 1 or 2 or 3

5. exp China/

6. (china or chinese or hubei or wuhan).af.

7. 5 or 6

8. 4 and 7

9. (betacoronavirus* or beta coronavirus* or coronavirus* or corona virus*).mp.

10. (severe acute respiratory syndrome coronavirus 2 or "SARS CoV-2" or "SARSCoV 2" or SARSCoV2 or cov2 or "sars 2" or COVID or "coronavirus 2" or covid19 or nCov or ((new or Novel) adj3 coronavirus*) or ncp).mp.

11. (exp pneumonia/ or pneumonia.mp.) and wuhan.af.

12. 8 or 9 or 10 or 11

13. quarantine/

14. social isolation/ or isolation/ or patient isolation/

15. loneliness/

16. psychology/

17. mental health/

18. mental disease/

19. social stigma/

20. fear/

21. anxiety/

22. depression/

23. physiological stress/ or mental stress/

24. anger/

25. irritability/

26. exp grief/

27. exp burnout/

28. (mental disorder* or Quarantine* or Self-isolation or isolation or social distanc* or shelter*-in-place or psych* or mental health or mental illness* or stigma or fear* or anxiety or anxious or depression or depressive or loneliness or stress* or trauma* or post-traumatic or posttraumatic or anger or mood* or irritability or irritable or emotional disturbance* or grief or burned out or burnout).tw,kw.

29. or/13-27

30. 12 and 29

31. ("20191231" or 2020* or 2021*).dc.

32. 30 and 31

PsycINFO (Ovid)

1. (coronavirus* or corona virus* or betacoronavirus* or beta coronavirus*).mp.

2. (severe acute respiratory syndrome coronavirus 2 or "SARS CoV-2" or "SARSCoV 2" or SARSCoV2 or cov2 or "sars 2" or COVID or "coronavirus 2" or covid19 or nCov or ((new or Novel) adj3 coronavirus*) or ncp).mp. or ((exp pneumonia/ or pneumonia.mp.) and wuhan.af.)

3. 1 or 2

4. ("20191231" or 2020* or 2021*).up.

5. 3 and 4

CINAHL

| **Search ID#** | **Search Terms** |
| --- | --- |
| S26 | S11 AND S25 |
| S25 | S12 OR S13 OR S14 OR S15 OR S16 OR S17 OR S18 OR S19 OR S20 OR S21 OR S22 OR S23 OR S24 |
| S24 | TI ( (mental disorder* or Quarantine* or Self-isolation or isolation or social distanc* or shelter*-in-place or psych* or mental health or mental illness* or stigma or fear* or anxiety or anxious or depression or depressive or loneliness or stress* or trauma* or post-traumatic or posttraumatic or anger or mood* or irritability or irritable or emotional disturbance* or grief or burned out or burnout) ) OR AB ( (mental disorder* or Quarantine* or Self-isolation or isolation or social distanc* or shelter*-in-place or psych* or mental health or mental illness* or stigma or fear* or anxiety or anxious or depression or depressive or loneliness or stress* or trauma* or post-traumatic or posttraumatic or anger or mood* or irritability or irritable or emotional disturbance* or grief or burned out or burnout) ) |
| S23 | (MH "Burnout, Professional") |
| S22 | (MH "Grief+") |
| S21 | (MH "Anger") |
| S20 | (MH "Stress, Physiological") OR (MH "Stress, Psychological") |
| S19 | (MH "Depression") |
| S18 | (MH "Anxiety") |
| S17 | (MH "Fear") |
| S16 | (MH "Stigma") |
| S15 | (MH "Mental Health") or (MH "Mental Disorders") |
| S14 | (MH "Psychology") |
| S13 | (MH "Social Isolation") OR (MH "Loneliness") or (MH “Social Distancing”) or (MH “Stay at Home Orders”) † |
| S12 | (MH "Quarantine") |
| S11 | S7 OR S8 OR S9 OR S10 |
| S10 | ( (MH "Pneumonia+") or TI (pneumonia) OR AB (pneumonia) ) AND ( TI (wuhan) OR AB (wuhan) OR AF (wuhan) ) |
| S9 | TI ( (severe acute respiratory syndrome coronavirus 2 or "SARS CoV-2" or "SARSCoV 2" or SARSCoV2 or cov2 or "sars 2" or COVID or "coronavirus 2" or covid19 or nCov or ((new or Novel) N3 coronavirus*) ) OR AB ( (severe acute respiratory syndrome coronavirus 2 or "SARS CoV-2" or "SARSCoV 2" or SARSCoV2 or cov2 or "sars 2" or COVID or "coronavirus 2" or covid19 or nCov or ((new or Novel) N3 coronavirus*) ) or (MH “Covid 19”) † |
| S8 | TI ( (betacoronavirus* or beta coronavirus* or coronavirus* or corona virus*) ) OR AB ( (betacoronavirus* or beta coronavirus* or coronavirus* or corona virus*)) |
| S7 | S5 AND S6 |
| S6 | S1 OR S2 |
| S5 | S3 OR S4 |
| S4 | TI ( (china or chinese or hubei or wuhan) ) OR AB ( (china or chinese or hubei or wuhan) ) OR AF ( (china or chinese or hubei or wuhan) ) OR SO ( (china or chinese or hubei or wuhan) ) |
| S3 | (MH "China+") |
| S2 | TI ( (betacoronavirus* or beta coronavirus* or coronavirus* or corona virus*) ) OR AB ( (betacoronavirus* or beta coronavirus* or coronavirus* or corona virus*) ) |
| S1 | (MH "Coronavirus+") OR (MH "Coronavirus Infections+") |

Web of Science

TOPIC: (Quarantine* or "Self-isolation" or isolation or "social distanc*" or "shelter*-in-place" or psych* or "mental health" or "mental illness*" or "mental disorder*" or stigma or fear* or anxiety or anxious or depression or depressive or loneliness or stress* or trauma* or "post-traumatic" or posttraumatic or anger or mood* or irritability or irritable or "emotional disturbance*" or grief or "burned out" or burnout) AND TOPIC: ((coronavirus* or "corona virus*" or betacoronavirus* or "beta coronavirus*" or "severe acute respiratory syndrome coronavirus 2" or "SARS CoV-2" or "SARSCoV 2" or SARSCoV2 or cov2 or "sars 2" or COVID or "coronavirus 2" or covid19 or nCov or "Novel coronavirus*" or "new coronavirus*"))

Indexes=SCI-EXPANDED, SSCI, A&HCI, CPCI-S, CPCI-SSH, BKCI-S, BKCI-SSH, ESCI, CCR-EXPANDED, IC Timespan=Year to date

China National Knowledge Infrastructure

Restricted to disciplines: Medical and Public Health & Social science

TI=(隔离+封城+社交距离+方舱+心理+心理健康+精神卫生+精神疾病+心理疾病+污名+耻辱+羞辱+恐惧+焦虑+抑郁+孤独+压力+应激+创伤+创伤后+愤怒+情绪+心情+易怒+情绪障碍+心理障碍+哀伤+悲伤+悲痛+悲哀+忧郁+倦怠)*(新冠+新型冠状) OR AB=(隔离+封城+社交距离+方舱+心理+心理健康+精神卫生+精神疾病+心理疾病+污名+耻辱+羞辱+恐惧+焦虑+抑郁+孤独+压力+应激+创伤+创伤后+愤怒+情绪+心情+易怒+情绪障碍+心理障碍+哀伤+悲伤+悲痛+悲哀+忧郁+倦怠)*(新冠+新型冠状)

Wanfang

题名:("隔离"+封城+"社交距离"+方舱+心理+"心理健康"+"精神卫生"+"精神疾病"+"心理疾病"+污名+耻辱+羞辱+恐惧+焦虑+抑郁+孤独+压力+应激+创伤+"创伤后"+愤怒+情绪+心情+易怒+”情绪障碍"+"心理障碍”+哀伤+悲伤+悲痛+悲哀+忧郁+倦怠)*("新冠"+"新型冠状")+摘要:("隔离"+封城+"社交距离"+方舱+心理+"心理健康"+"精神卫生"+"精神疾病"+"心理疾病"+污名+耻辱+羞辱+恐惧+焦虑+抑郁+孤独+压力+应激+创伤+"创伤后"+愤怒+情绪+心情+易怒+"情绪障碍"+"心理障碍"+哀伤+悲伤+悲痛+悲哀+忧郁+倦怠)*("新冠"+"新型冠状")

We made several amendments to the original search strategies. Since the Wanfang database cannot export more than 5000 references at once, we broke the search strategies into two or more smaller search strings to get all the references. The four changes on September 1, 2020, September 28, 2020, October 15, 2020 and October 18, 2020 are all for this purpose.

To make this process more efficient, the disciplines of the China National Knowledge Infrastructure database were restricted to Medical and Public Health AND Social science subgroup 2 and those of Wanfang database were restricted to Medicine and Health AND Culture, Science, Education and PE disciplines on October 23, 2020.

September 1, 2020

Wanfang

题名:("隔离"+封城+"社交距离"+方舱+心理+"心理健康"+"精神卫生"+"精神疾病"+"心理疾病"+污名+耻辱+羞辱+恐惧+焦虑)*("新冠"+"新型冠状")+摘要:("隔离"+封城+"社交距离"+方舱+心理+"心理健康"+"精神卫生"+"精神疾病"+"心理疾病"+污名+耻辱+羞辱+恐惧+焦虑)*("新冠"+"新型冠状")

题名:("隔离"+封城+"社交距离"+方舱+抑郁+孤独+压力+应激+创伤+"创伤后"+愤怒+情绪+心情+易怒+"情绪障碍"+"心理障碍"+哀伤+悲伤+悲痛+悲哀+忧郁+倦怠)*("新冠"+"新型冠状")+摘要:("隔离"+封城+"社交距离"+方舱+抑郁+孤独+压力+应激+创伤+"创伤后"+愤怒+情绪+心情+易怒+"情绪障碍"+"心理障碍"+哀伤+悲伤+悲痛+悲哀+忧郁+倦怠)*("新冠"+"新型冠状")

September 28, 2020

Wanfang

题名:("隔离"+封城+"社交距离"+方舱+心理+"心理健康"+"精神卫生"+"精神疾病"+"心理疾病"+污名+耻辱+羞辱+恐惧+焦虑)*("新冠"+"新型冠状")+摘要:("隔离"+封城+"社交距离"+方舱+心理+"心理健康"+"精神卫生"+"精神疾病"+"心理疾病"+污名+耻辱+羞辱+恐惧+焦虑)*("新冠"+"新型冠状")

题名:("隔离"+封城+"社交距离"+方舱+抑郁+孤独+压力+应激+创伤+"创伤后")*("新冠"+"新型冠状")+摘要:("隔离"+封城+"社交距离"+方舱+抑郁+孤独+压力+应激+创伤+"创伤后")*("新冠"+"新型冠状")

题名:("隔离"+封城+"社交距离"+方舱+愤怒+情绪+心情+易怒+"情绪障碍"+"心理障碍"+哀伤+悲伤+悲痛+悲哀+忧郁+倦怠)*("新冠"+"新型冠状")+摘要:("隔离"+封城+"社交距离"+方舱+愤怒+情绪+心情+易怒+"情绪障碍"+"心理障碍"+哀伤+悲伤+悲痛+悲哀+忧郁+倦怠)*("新冠"+"新型冠状")

October 15, 2020

Wanfang

题名:("隔离"+封城+"社交距离"+方舱+心理+"心理健康"+"精神卫生"+"精神疾病"+"心理疾病")*("新冠"+"新型冠状")+摘要:("隔离"+封城+"社交距离"+方舱+心理+"心理健康"+"精神卫生"+"精神疾病"+"心理疾病")*("新冠"+"新型冠状")

题名:("隔离"+封城+"社交距离"+方舱+污名+耻辱+羞辱+恐惧+焦虑+抑郁+孤独+压力)*("新冠"+"新型冠状")+摘要:("隔离"+封城+"社交距离"+方舱+污名+耻辱+羞辱+恐惧+焦虑+抑郁+孤独+压力)*("新冠"+"新型冠状")

题名:("隔离"+封城+"社交距离"+方舱+应激+创伤+"创伤后"+愤怒+情绪+心情+易怒+"情绪障碍"+"心理障碍"+哀伤+悲伤+悲痛+悲哀+忧郁+倦怠)*("新冠"+"新型冠状")+摘要:("隔离"+封城+"社交距离"+方舱+应激+创伤+"创伤后"+愤怒+情绪+心情+易怒+"情绪障碍"+"心理障碍"+哀伤+悲伤+悲痛+悲哀+忧郁+倦怠)*("新冠"+"新型冠状")

October 18, 2020

Wanfang

题名:("隔离"+封城+"社交距离"+方舱+心理+"心理健康"+"精神卫生"+"精神疾病"+"心理疾病")*("新冠"+"新型冠状")+摘要:("隔离"+封城+"社交距离"+方舱+心理+"心理健康"+"精神卫生"+"精神疾病"+"心理疾病")*("新冠"+"新型冠状")

题名:("隔离"+封城+"社交距离"+方舱+污名+耻辱+羞辱+恐惧+焦虑+抑郁)*("新冠"+"新型冠状")+摘要:("隔离"+封城+"社交距离"+方舱+污名+耻辱+羞辱+恐惧+焦虑+抑郁)*("新冠"+"新型冠状")

题名:("隔离"+封城+"社交距离"+方舱+孤独+压力)*("新冠"+"新型冠状")+摘要:("隔离"+封城+"社交距离"+方舱+孤独+压力)*("新冠"+"新型冠状")

题名:("隔离"+封城+"社交距离"+方舱+应激+创伤+"创伤后"+愤怒+情绪+心情+易怒+"情绪障碍"+"心理障碍"+哀伤+悲伤+悲痛+悲哀+忧郁+倦怠)*("新冠"+"新型冠状")+摘要:("隔离"+封城+"社交距离"+方舱+应激+创伤+"创伤后"+愤怒+情绪+心情+易怒+"情绪障碍"+"心理障碍"+哀伤+悲伤+悲痛+悲哀+忧郁+倦怠)*("新冠"+"新型冠状")

October 23, 2020

China National Knowledge Infrastructure

Restricted to disciplines: Medical and Public Health & Social science subgroup 2

TI=(隔离+封城+社交距离+方舱+心理+心理健康+精神卫生+精神疾病+心理疾病+污名+耻辱+羞辱+恐惧+焦虑+抑郁+孤独+压力+应激+创伤+创伤后+愤怒+情绪+心情+易怒+情绪障碍+心理障碍+哀伤+悲伤+悲痛+悲哀+忧郁+倦怠)*(新冠+新型冠状) OR AB=(隔离+封城+社交距离+方舱+心理+心理健康+精神卫生+精神疾病+心理疾病+污名+耻辱+羞辱+恐惧+焦虑+抑郁+孤独+压力+应激+创伤+创伤后+愤怒+情绪+心情+易怒+情绪障碍+心理障碍+哀伤+悲伤+悲痛+悲哀+忧郁+倦怠)*(新冠+新型冠状)

Wanfang

Restricted to disciplines: Medicine and Health & Culture, Science, Education and PE

题名:("隔离" or 封城 or "社交距离" or 方舱 or 心理 or "心理健康" or "精神卫生" or "精神疾病" or "心理疾病" or 污名 or 耻辱 or 羞辱 or 恐惧 or 焦虑 or 抑郁 or 孤独 or 压力 or 应激 or 创伤 or "创伤后" or 愤怒 or 情绪 or 心情 or 易怒 or "情绪障碍" or "心理障碍" or 哀伤 or 悲伤 or 悲痛 or 悲哀 or 忧郁 or 倦怠) and ("新冠" or "新型冠状") or 摘要:("隔离" or 封城 or "社交距离" or 方舱 or 心理 or "心理健康" or "精神卫生" or "精神疾病" or "心理疾病" or 污名 or 耻辱 or 羞辱 or 恐惧 or 焦虑 or 抑郁 or 孤独 or 压力 or 应激 or 创伤 or "创伤后" or 愤怒 or 情绪 or 心情 or 易怒 or "情绪障碍" or "心理障碍" or 哀伤 or 悲伤 or 悲痛 or 悲哀 or 忧郁 or 倦怠) and ("新冠" or "新型冠状").

MedRxiv (pre-prints)

Search 1: (isolation OR “mental health” OR “mental illness” OR “mental disorder”) AND (COVID OR covid19)

Search 2: (psychology OR psychological OR psychosocial OR anxiety OR depression OR stress or trauma) AND (COVID OR covid19)

Open Science Framework (pre-prints)

(isolation OR psychology OR psychological OR psychosocial OR “mental health” OR “mental illness” OR “mental disorder” OR anxiety OR depression OR stress or trauma) AND (coronavirus OR COVID OR covid19)

**Supplemental Material 2.** Title and Abstract and Full-Text Review Inclusion and Exclusion Criteria Coding Guides

Title and Abstract Coding Criteria

**No: not original human data or a case study or case series.** If it is clear from the title and abstract that the article is not an original report of primary data, but, for example, a letter, editorial, systematic review or meta-analysis, or it is a single case study or case series, then it is excluded. Studies reporting only on animal, cellular, or genetic data are also excluded. Conference abstracts are included.

**No: not a study of any population affected by the COVID-19 outbreak.** If it is clear from the title or abstract that the study is not about any population affected by the COVID-19 outbreak, it is excluded. Studies that include fewer than 100 subjects, are excluded. If a longitudinal study has baseline sample size with at least 100 participants, but no follow-up with at least 100 participants, then we exclude the study (and document); if its baseline and at least one follow-up have more than 100 participants, we include the study.

**No: not a study which reports mental health symptom changes longitudinally pre-COVID-19 to COVID-19 or during COVID-19.** If it is clear from the title or abstract that the study does not report continuous scores of symptom levels or proportions of participants meeting the threshold on a validated scale, or diagnostic criteria using a validated diagnostic interview prior to and after the start of COVID-19, or longitudinally during COVID-19, then it will be excluded.

For pre-COVID versus during-COVID studies, pre- and during- samples must include the same cohort, not different representative samples. Pre- and during-samples should have less than 10% difference in the participants in the sample* or should statistically account for missing data, i.e., if N between the samples differs by more than 10%, modelling or imputation is needed to evaluate results for all participants. Pre-COVID data needs to be collected prior to 2020 (or at least 80% of the participants’ data need be collected prior to 2020 if collection spans from 2019 to 2020) and after 2018 (or at least 80% of the participants’ data need to be collected after 2018 if collection spans from pre-2018 to 2018).

For studies with multiple waves across COVID, if there are pre-pandemic time points, the most recent pre-pandemic wave needs to be in 2018 or later; if the most recent pre-pandemic wave spans from pre-2018 to 2018, at least 80% of the data need to be collected in 2018. Studies with multiple waves across COVID-19 must have at least two time points that have less than 10% difference in the participants in the sample*, or should statistically account for missing data, regardless of whether or not the study has pre-COVID assessments.

If outcomes from the study are only shown graphically without eligible numerical values, exclude the study.

* At least 90% of participants in assessments from two time points need to be the same participants. In a three-wave survey, if N-T1 = 1000, N – T2 = 500, and N – T3 = 500, T2 and T3 would only be eligible if at least 90% of the participants at each time point were the same. It is not enough to just have a total N within 10%.

**Yes: study eligible for inclusion in full-text review.**

Full Text Coding Criteria

**No: not original human data or a case study or case series.** If the article is not an original report of primary data, but, for example, a letter, editorial, systematic review or meta-analysis, or it is a single case study or case series, then it is excluded. Studies reporting only on animal, cellular, or genetic data are also excluded. Conference abstracts are included.

**No: not a study of any population affected by the COVID-19 outbreak.** If the study is not about any population affected by the COVID-19 outbreak, it is excluded. Studies that include fewer than 100 subjects, are excluded. If a longitudinal study has baseline sample size with at least 100 participants, but no follow-up with at least 100 participants, then we exclude the study (and document); if its baseline and at least one follow-up have more than 100 participants, we include the study.

**No: not a study which reports mental health symptom changes longitudinally pre-COVID-19 to COVID-19 or during COVID-19.** If the study does not report continuous scores of symptom levels or proportions of participants meeting the threshold on a validated scale, or diagnostic criteria using a validated diagnostic interview prior to and after the start of COVID-19, or longitudinally during COVID-19, then it will be excluded.

For pre-COVID versus during-COVID studies, pre- and during- samples must include the same cohort, not different representative samples. Pre- and during-samples should have less than 10% difference in the participants in the sample* or should statistically account for missing data, i.e., if N between the samples differs by more than 10%, modelling or imputation is needed to evaluate results for all participants. Pre-COVID data needs to be collected prior to 2020 (or at least 80% of the participants’ data need be collected prior to 2020 if collection spans from 2019 to 2020) and after 2018 (or at least 80% of the participants’ data need to be collected after 2018 if collection spans from pre-2018 to 2018).

For studies with multiple waves across COVID, if there are pre-pandemic time points, the most recent pre-pandemic wave needs to be in 2018 or later; if the most recent pre-pandemic wave spans from pre-2018 to 2018, at least 80% of the data need to be collected in 2018. Studies with multiple waves across COVID-19 must have at least two time points that have less than 10% difference in the participants in the sample*, or should statistically account for missing data, regardless of whether or not the study has pre-COVID assessments.

If outcomes from the study are only shown graphically without eligible numerical values, exclude the study.

* At least 90% of participants in assessments from two time points need to be the same participants. In a three-wave survey, if N-T1 = 1000, N – T2 = 500, and N – T3 = 500, T2 and T3 would only be eligible if at least 90% of the participants at each time point were the same. It is not enough to just have a total N within 10%.

**Yes: study eligible for inclusion in systematic review.**

**Supplemental Material 3.** Adequacy of Study Methods and Reporting – Adapted Joanna Briggs Institute Checklist for Prevalence Studies

**Q1. Was the sample frame appropriate to address the target population?**

**Yes:** The sampling frame was a true or close representation of the target population.

**No:** The sampling frame was NOT a true or close representation of the target population.

**Unclear:** Not enough information provided to determine.

**Q2. Were study participants recruited in an appropriate way?**

**Yes:** A census was undertaken, OR, some form of random selection was used to select the sample (e.g. simple random sampling, stratified random sampling, cluster sampling, systematic sampling).

**No:** A census was NOT undertaken, AND some form of random selection was NOT used to select the sample.

**Unclear:** Not enough information provided to determine.

**Q3. Was the sample size adequate?**

**Yes:** There is evidence that the authors conducted a sample size calculation to determine an adequate sample size OR the study was large enough (e.g., a large national survey) whereby a sample size calculation is not required. In these cases, sample size can be considered adequate. If at least 200 participants are included for continuous outcomes and 250 for proportions, this is considered low risk.

**No:** The authors did not reach their intended sample size, or no sample size calculation is provided and there are < 100 participants for continuous outcomes, or < 125 for proportions.

**Unclear:** No sample size calculation is provided, and between 100-199 participants are included for continuous outcomes or between 125-249 for proportions.

**Q4. Were the study participants and setting described in detail?**

**Yes:** Data included age, sex, and at least 1 socioeconomic indicator (e.g., income, education, work status).

**No:** The minimum sociodemographic variables have not been reported.

**Unclear:** Not stated

**Q5. Was the response rate adequate and was the data analysis conducted with sufficient coverage?**

**Yes:** The overall response rate or response rate for intended subgroups was >/=75%, OR, an analysis was performed that established that there was not a substantive difference in relevant demographic characteristics between responders and non-responders within a subgroup (if non-response too high (e.g., > 50%), code “No”)

**No:** The overall response rate or response rate for subgroups was <75%, and if any analysis comparing responders and non-responders was done, it showed a meaningful difference in relevant demographic characteristics between responders and non-responders.

**Unclear:** Not enough information provided to determine.

**Q6. Were valid methods used for the identification of the outcome variable?**

**Yes:** The study instrument had been shown to have reliability and validity, e.g., test-retest, piloting, validation in a previous study, etc.

**No:** The study instrument had NOT been shown to have reliability or validity.

**Unclear:** Not stated.

**Q7. Was the mental health outcome measured in a standard, reliable way for all participants?**

**Yes:** All self-report data were collected directly from the participants. Any clinical interview data includes at least information about the interviewers’ level of education or training received. The same mode of data collection was used for all participants. All aspects of this question must be present (where relevant).

**No:** In some instances, data were collected from a proxy (e.g., a spouse). The qualifications of clinical interviewers are not reported or not appropriate. The same mode of data collection was NOT used for all participants. If any aspects of this item are absent, it is high risk.

**Unclear**: Not stated.

**Q8. Was there appropriate statistical analysis?**

**Yes:** Continuous variables report (1) mean (SD) of change or (2) pre mean (SD) and post mean (SD) with/out correlation between pre and post scores. For dichotomous variables, numerator, denominator, and percentages are clearly reported. Continuous variables are not artificially dichotomized. The statistical analyses section is detailed enough for readers to understand change scores (see STROBE reporting guidelines, if necessary).

**No:** Continuous variables do not include a report of the (1) mean (SD) of change or (2) pre mean (SD) and post mean (SD) with/out correlation between pre and post scores. For dichotomous variables, the numerator, denominator, or percentages are not clearly reported. The statistical analyses section does not clearly describe the methods used to assess change scores.

**Q9. Was the follow-up rate adequate, and if not, was the low follow-up rate managed appropriately?**

**Yes:** At least 75% of those who participated in the pre-COVID-19 assessment(s) provided follow-up responses and had their responses included in the follow-up, OR, an analysis was performed that showed no substantive difference in relevant demographic characteristics between participants who stayed in the study and drop-outs (if dropout too high (e.g. > 50%), code “No”).

**No:** Less than 75% of those participated in the pre-COVID-19 assessment(s) provided responses and had their responses included in the follow-up, and if any analysis comparing participants who stayed in the study and drop-outs was done, it showed a substantive difference in relevant demographic characteristics between the two groups.

**Unclear:** Not stated.

**Supplementary Table 1.** Confidence Intervals for Proportion Change When Assuming that 30% and 70% of Pre-COVID-19 Cases were Cases During COVID-19^a^

| **First Author** | **Outcome Domain** | **Sex/Gender** | **95% CI with 30%** | **95% CI with 70%** |
| --- | --- | --- | --- | --- |
| Dongb | General Mental Health | Females/Women | -0.10, -0.06 | -0.10, -0.07 |
|  |  | Males/Men | -0.10, -0.04 | -0.10, -0.04 |
| Saraswathi^b^ | Anxiety Symptoms | Females/Women | -0.24, -0.03 | -0.22, -0.06 |
|  |  | Males/Men | -0.23, 0.06 | -0.20, 0.02 |
|  | Depression Symptoms | Females/Women | -0.10, 0.14 | -0.05, 0.09 |
|  |  | Males/Men | -0.25, 0.05 | -0.21, 0.01 |
|  | Stress | Females/Women | -0.12, 0.06 | -0.09, 0.04 |
|  |  | Males/Men | -0.20, 0.08 | -0.16, 0.03 |
| Sigursteinsdottir | Stress | Females/Women | -0.14, -0.04 | -0.10. 1.04 |
|  |  | Males/Men | -0.19, -0.01 | -0.13, -0.08 |
| van der Velden^b^ | General Mental Health | Females/Women | -0.02, 0.03 | -0.01, 0.02 |
|  |  | Males/Men | -0.03, 0.01 | -0.02, 0.00 |
| van der Velden^b^ | General Mental Health | Females/Women | -0.01, 0.03 | -0.00, 0.03 |
|  |  | Males/Men | -0.03, 0.01 | -0.03, 0.00 |
| Yu | General Mental Health | Females/Women | -0.11, -0.00 | -0.06, -0.05 |
|  |  | Males/Men | -0.12, -0.04 | -0.083, -0.075 |

^a^Values calculated for studies that did not provide 95% confidence intervals for change. ^b^Results included in the previous review (Dal Santo et al.^2^).

**Supplementary Figure 1.** PRISMA Flowchart

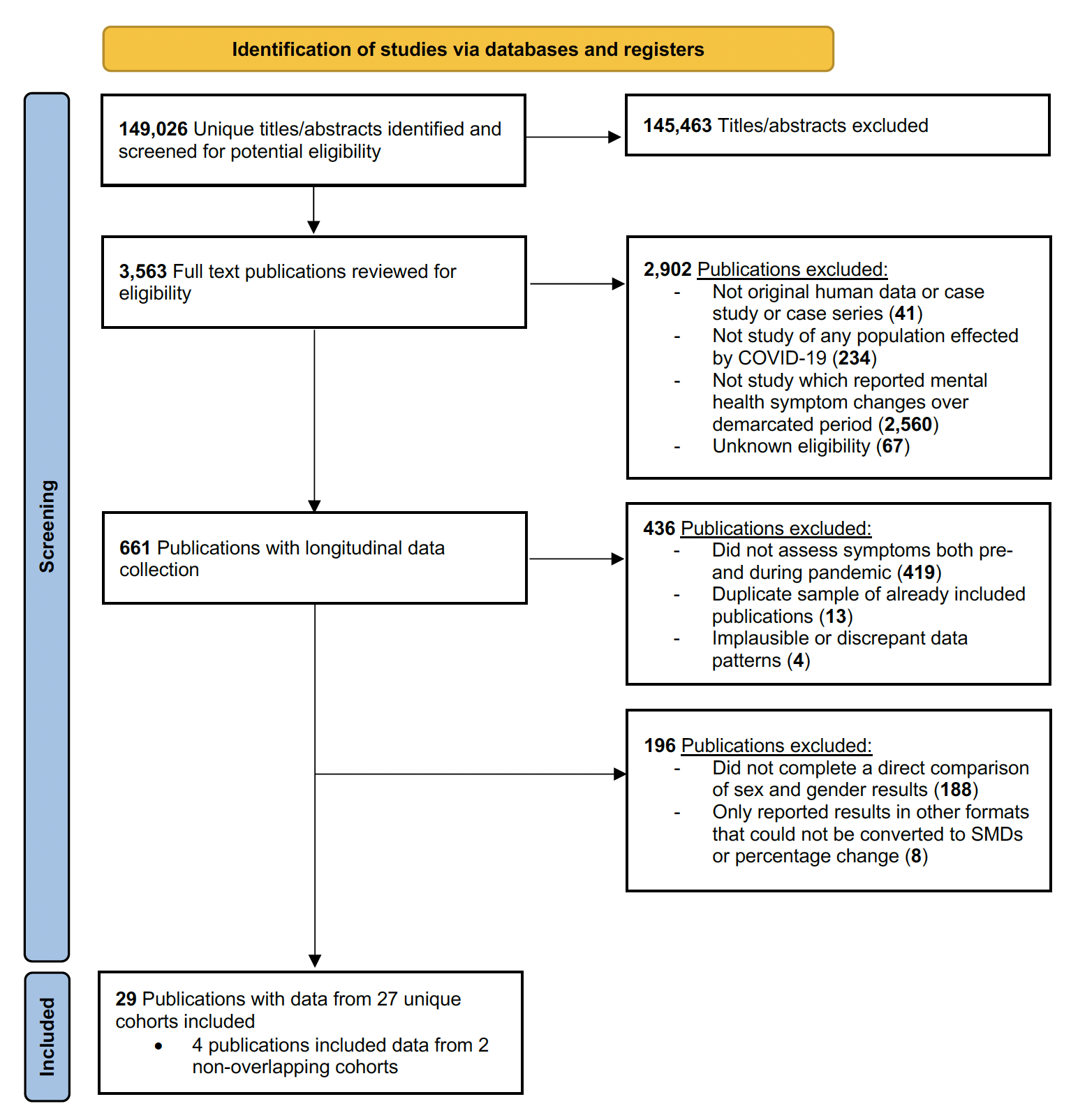

**Supplementary Table 2.** Adequacy of Methods and Reporting of Included Studies (N=29)

| **Author** | **Appropriate**  **sample**  **frame** | **Appropriate participant recruitment** | **Adequate**  **sample size** | **Subjects**  **and setting adequately described** | **Adequate response rate and data analysis with sufficient coverage** | **Valid methods for identification of outcome variable** | **Standard, reliable outcome measurement** | **Appropriate statistical analysis** | **Adequate follow-up response rate/ appropriate management of low response rate** |
| --- | --- | --- | --- | --- | --- | --- | --- | --- | --- |
| Adesogan | No | No | Yes | Yes | Unclear | Yes | No | Yes | Yes |
| Cerniglia | No | Unclear | Yes | Yes | Unclear | Yes | Yes | Yes | Yes |
| Cheung | No | Unclear | Yes | Yes | Unclear | Yes | Yes | Yes | Unclear |
| Dong^a^ | No | No | Yes | Yes | Yes | Yes | Yes | Yes | Yes |
| Frank | No | Unclear | Unclear | Yes | Unclear | Yes | Yes | Yes | Unclear |
| He | No | Yes | Yes | Yes | Yes | Yes | Yes | Yes | Yes |
| Kumar | Yes | Unclear | Yes | Yes | Unclear | Yes | No | Yes | Unclear |
| Li | No | Unclear | Yes | Yes | Unclear | Yes | Yes | Yes | Yes |
| Lim^a^ | Yes | Unclear | Yes | Yes | Unclear | Yes | Unclear | Yes | Unclear |
| Lucassen | No | No | Unclear | No | Unclear | Yes | Yes | Yes | Yes |
| Magson^a^ | No | Unclear | Yes | Yes | Unclear | Yes | Yes | Yes | No |
| Megías-Robles^a^ | Unclear | No | Unclear | No | Unclear | Yes | Yes | Yes | No |
| Mei | No | Unclear | Yes | Yes | Unclear | Yes | Yes | Yes | No |
| Meireles | No | Unclear | Yes | Yes | Unclear | Yes | Yes | Yes | Unclear |
| Pierce^a^ | Yes | Yes | Yes | Yes | Unclear | Yes | Yes | Yes | Unclear |
| Daly^a^ |  |  |  |  |  |  |  |  | No |
| Pedrini | No | No | Unclear | Yes | Unclear | Yes | Unclear | Yes | Yes |
| Rimfield^a^ | Yes | Unclear | Yes | Yes | Unclear | Yes | Yes | Yes | Unclear |
| Rohde | No | Unclear | Yes | Yes | No | Yes | Yes | Yes | No |
| Saraswathi^a^ | No | Yes | Unclear | Yes | Yes | Yes | Yes | Yes | Yes |
| Savage^a^ | No | Yes | Yes | Yes | No | Yes | Yes | Yes | No |
| Shanahan^a^ | No | Unclear | Yes | Yes | Unclear | Yes | Yes | Yes | Yes |
| Shuval | No | No | Yes | Yes | Unclear | Yes | Yes | Yes | Unclear |
| Sigursteinsdottir | No | Unclear | Yes | Yes | Unclear | Yes | Yes | Yes | No |
| Soltanzadeh | No | Yes | Yes | Yes | Yes | Yes | Unclear | Yes | Yes |
| van der Velden^a^ | Yes | Yes | Yes | Yes | Yes | Yes | Yes | Yes | Yes |
| van der Velden^a^ |  |  |  |  |  |  |  |  |  |
| Yu | No | No | Yes | No | Yes | Yes | Yes | Yes | Yes |
| Zhang | Yes | Unclear | Yes | Yes | Unclear | Yes | Yes | Yes | No |

^a^Results included in the previous review (Dal Santo et al.^2^).
